# Response of human liver tissue to innate immune stimuli

**DOI:** 10.1101/2020.12.22.20248755

**Authors:** Xia Wu, Jessica B. Roberto, Allison Knupp, Alexander L. Greninger, Camtu D. Truong, Nicole Hollingshead, Heidi L. Kenerson, Marianne Tuefferd, Antony Chen, Helen Horton, Keith R. Jerome, Stephen J. Polyak, Raymond S. Yeung, Ian N. Crispe

## Abstract

Precision-cut human liver slice cultures (PCLS) has become an important alternative immunological platform in preclinical testing. To further evaluate the capacity of PCLS, we investigated the innate immune response to TLR3 agonist (poly-I:C) and TLR4 agonist (LPS) with the normal and pathological liver tissue. Pathological liver tissue was obtained from patients with active chronic HCV infection, and patients with former chronic HCV infection cured by recent Direct-Acting Antiviral (DAA) drug therapy. We found that hepatic innate immunity was not suppressed but enhanced in the HCV-infected tissue, compared with the healthy controls. Furthermore, despite recent HCV elimination, DAA-cured liver tissue manifested ongoing abnormalities in liver immunity. Sustained abnormal immune gene expression in DAA-cured samples were identified from direct *ex vivo* measurements and with TLR3 and TLR4 stimulation assays. Those genes that were up-regulated in chronic HCV-infected liver tissue were enriched in expression in the liver non-parenchymal cell compartment. These results demonstrated the utility of PCLS in studying the liver pathology and innate immunity.

## Introduction

We recently reported the development of human liver slice technology (PCLS) as an immunological platform^1,2^. Compared with other liver research methods, the PCLS method has the advantage that hepatocytes and the major subsets of non-parenchymal cells (KC, LSEC and HSC) are cultured together in their normal anatomical relationships, enabling the analysis of liver cell function in the context of intercellular interaction. Furthermore, the patterns of liver disease that are present in fresh tissue could be maintained over time in the slice cultures^1^. For example, steatotic liver slices retains fat droplets, and cholestatic liver slices were manifested with yellow-green pigment in hepatocytes. However, it remained unclear whether PCLS has the capacity to reveal the dynamic alternations in innate immune response that may occur in pathological liver tissue compared with healthy controls. As a proof of principle, we compared the dynamic response with PCLS with liver tissue collected from three groups of patients which included non-infected subjects, chronic HCV-infected patients, and patients whose HCV was cured by DAA treatment.

HCV-infected liver tissue is of particular interest, because HCV infection is one of the most common liver diseases in the USA and globally. Approximately 71 million individuals are infected with HCV worldwide ^3^. The acute phase of infection is often subclinical, but 55–85% of infected individuals develop a chronic infection leading to progressive liver pathology ^4^. The World Health Organization (WHO) estimated that approximately 399,000 people died from HCV-related diseases in 2016, mostly from cirrhosis and hepatocellular carcinoma (HCC).

The recent introduction of highly effective direct-acting antiviral (DAA) drugs has presented a unique opportunity to investigate the resolution of pathological liver to normality after HCV elimination. The extent to which such drugs revert the liver to normal is not fully resolved, with some studies suggesting that HCV clearance with DAA therapy leaves residual abnormalities in innate immunity in peripheral monocytes and NK cells ^5,6^, adaptive immunity in peripheral CD4+ and CD8+ T cells ^7-10^, in γδ-T cells ^11^, in mucosal associated invariant T cells ^12^, and in the persisting risk to hepatocellular cancer ^13-16^. Other studies also found abnormal serum lipids ^17^, persistent epigenetic modifications ^18,19^, and sustained hepatic inflammation ^20^ in liver biopsies after DAA treatment. To our knowledge, no studies have evaluated hepatic immunity of untreated versus DAA-treated HCV infection in the context of intact liver tissue.

## Materials and Methods

### Liver samples, preparation and culturing

Fresh non-tumor liver tissues were obtained from patients undergoing liver resection at the University of Washington Medical Center (Seattle, WA, USA). All patients in this study prospectively consented to donate liver tissue for research under the Institutional Review Board protocol #00001852. Tissue from DAA-treated patients were collected from those who achieved sustained virologic response (SVR), with HCV viral load being non-detected after the completion of DAA therapy. Clinical details of these patients are provided in **Table S1**.

Liver slicing and culturing were as previously described ^1^. Briefly, liver cores of 6 mm diameter were excised from the resected liver tissue using a biopsy punch (Integra Miltex, York, PA, USA), stored in BELZER-UW solution (Bridge to Life Ltd., Columbia, SC, USA), and transferred to the research laboratories typically within 1 h of tissue excision. Slices 250 μm thick were prepared using a vibrating microtome, Leica VT1200 S (Nussloch, Germany), using Dulbecco’s Modified Eagles Medium (DMEM) as the cutting medium. Liver slices were cultured individually on 0.4 µm millicell organotypic inserts in 24 well plates (Millipore Corporation, Billerica, MA, USA). The culturing medium comprised 1 × advanced-DMEM medium, 5% Fetal Bovine Serum (FBS), 1 × GlutaMAX, 0.5 × Penicillin-Streptomycin, 1 × Insulin-Transferrin-Selenium supplement and 15 mM HEPES (pH 7.2 - 7.5) (all from Gibco, Grand Island, NY, USA). The liver cultures were maintained on a rocking platform at 17 rpm in a humidified incubator at 5% CO_2_ and atmospheric concentration of O_2_ at 37°C. The medium was renewed every two to three days.

### Histology of liver slices

Liver slices were fixed with the 10% neutral-buffered formalin at room temperature for 24 h. The fixed liver slices were embedded in paraffin and were sliced into 4 μm-thick sections. Trichrome stain, picrosirius red stain, and hematoxylin and eosin (H&E) stain were analyzed with the standard protocol in the Pathology Research Services Laboratory at Department of Laboratory Medicine and Pathology at UW. The fibrosis score of liver slices was analyzed by a pathologist blinded from the patient clinical diseases. The Scheuer/Batts-Ludwig method was used for fibrosis scoring, with 0: No fibrosis, 1: portal fibrosis, 2: peri-portal fibrosis, 3: bridging fibrosis, and 4: cirrhosis. The whole-slide images were recorded with Nanozoomer Whole Slide Scanner (Hamamatsu City, Shizuoka Pref., Japan) and visualized with NDP.view2 software (Hamamatsu).

### Liver perfusion and isolation of liver cells

Human liver cell isolation procedures were adapted from several sources ^21,22^. Cell isolation was performed on wedges of resected tissue rather than cores. Perfusion buffer contained 1 × Hank’s Balanced Salt Solution (HBSS, without Ca++, Mg++, or phenol red, from Gibco), 10 mM HEPES (pH 7.2-7.5) and 0.5 mM EDTA (pH 8.0). Collagenase buffer contained 1 × HBSS (Gibco), 5 mM MgCl_2_, 5 mM CaCl_2_, 5 mM HEPES (pH 7.2-7.5), 0.5% w/v Collagenase IV (Sigma-Aldrich, St. Louis, MO, USA), 0.25% w/v Protease (Sigma), 0.125% w/v Hyaluronidase (Sigma), 0.05% w/v DNase I (Sigma). Fresh aliquots of enzymes were added to the buffer on the day of the perfusion experiment. Forty mL of Perfusion buffer and 20 mL of Collagenase buffer were used for each 10 g of liver tissue. Buffers were pre-warmed to 37°C prior to the perfusion step.

Liver tissue wedges were sequentially perfused with Washing buffer (1 × HBSS and 10 mM HEPES, pH 7.2-7.5), Perfusion buffer, Washing buffer, and the recirculating Collagenase buffer at a flow rate of 12 mL/min (Gilson’s MINIPULS 3, Middleton, WI, USA). The perfused liver tissue samples were gently mashed with a sterile syringe plunger through a sterile mesh strainer in ice-cold DMEM medium. Cell extracts were filtered through a 100 μm sterile strainer and centrifuged at 50 × *g* at 4°C for 3 min to enrich for hepatocytes in the pellets and non-parenchymal cells in the supernatant. The supernatants were transferred to a new tube and kept on ice. The pellets were washed three times with ice-cold DMEM medium, and were pelleted each time at 50 × *g* at 4°C for 3 min. To further improve the purity of the isolated hepatocytes, cell pellets were resuspended with 5 mL of ice-cold PBS, overlaid with 10 mL of 25% Percoll gradient solution. The mixture was centrifuged at 1400 × *g* (no brake) at 4°C for 20 min. The pellet contained the purified live hepatocytes. The viability of the isolated hepatocytes was determined with the trypan blue exclusion assay (Thermo Fisher Scientific, Waltham, MA, USA). If the viability was greater than 50%, the isolated hepatocytes were stored for RNA extraction.

For the non-parenchymal cells (NPC), the 50 x *g* centrifuge supernatants were further centrifuged at 500 × *g* at 4°C for 7 min. The pellets were resuspended in 5 mL of ice-cold PBS, and overlaid on top with 50% and 25% Percoll gradients (10 mL layers each), and were centrifuged at 1400 × *g* (no brake) at 4°C for 20 min. Cell layers were collected into 20 mL PBS each, and centrifuged again at 500 × *g* at 4°C for 7 min. A fraction of the pellets were saved for RNA extraction, which were the total NPC. The rest of the pellets were resuspended with the ice-cold Flow buffer containing 1 × PBS, 2% FBS, and 1 mM EDTA. Experimental Overview is illustrated in Fig. S1.

### Flow cytometry and cell sorting

The isolated non-parenchymal cells were stained with an antibody mixture that included eBioscience (San Diego, CA, USA): anti-CD45 (Cat. No. 15-0459-42); BioLegend (San Diego, CA, USA): anti-CD3 (Cat. No. 317330), anti-CD11b (Cat. No. 553310), anti-CD14 (Cat. No. 301834), anti-CD31 (Cat. No. 303120), anti-CD32 (Cat. No. 303206), anti-CD68 (Cat. No. 333814), and anti-CD271 (Cat. No. 345110). In addition, cells were also stained with LIVE/DEAD Fixable Far Red Dead Cell Stain Kit (Cat. No. L10120, Life Technologies, Carlsbad, CA, USA). The incubation mixture was kept at 4°C for 30 min in the dark on a rocking platform. The mixture was centrifuged with 500 × *g* at 4°C for 7 min. Cells were washed once with Flow buffer.

The antibody-labeled cells were sorted with a BD Aria III (BD Biosciences, San Jose, CA, USA). Analysis of cell populations was performed using FlowJo software, version 9.8.5 (FlowJo, LLC, Ashland, OR, USA). Kupffer cells were selected as the CD45+, CD3−, CD14+, CD68+, CD32+ populations ^23,24^. LSECs were selected as CD45−, CD31+, CD11b− ^25^. HSCs were selected as CD45−, CD271+, autofluorescence positive with the emission wavelength at 460 nm, SSC-H > 150 units ^26,27^ (**Fig. S1**). Cells were pelleted at 500 × *g* at 4°C for 7 min, and were stored in -80°C before RNA extraction.

### RNA isolation and qRT-PCR analysis

The RNA of liver slices or the purified liver cells was isolated with TRIzol and the Direct-zol RNA MiniPrep Kit (Zymo Research, Irvine, CA, USA). The RNA concentration was measured with nanoDrop (Thermo Fisher Scientific). The cDNA was synthesized with the QuantiTect Reverse Transcription Kit (Qiagen, Hilden, Germany).

A pre-amplification was performed before the multiplex qRT-PCR assays to improve the detection sensitivity ^28,29^, which included PCR reactions of cDNA as templates, and the 0.2 fold-diluted primer mixture of TaqMan assays of interest (Thermo Fisher Scientific) and the BIO-X-ACT Short Mix reagents (Bioline USA Inc., Taunton, MA, USA). The pre-amplified samples were diluted five-fold with RNase- and DNase-free H_2_O. Samples were analyzed with a 48 × 48 dynamic array and a BioMark HD microfluidics system (Fluidigm, San Francisco, CA, USA). The Fluidigm Real-Time PCR Analysis software was used to calculate Ct thresholds, using the settings of quality threshold 0.65, baseline correction linear, Ct threshold method auto detection. Gene abundance of individual liver slices was normalized to the arithmetic mean of Ct values of ACTB, HPRT, and GAPDH. Typically, three liver slices were analyzed for each subject for each time point. The arithmetic mean delta Ct of three liver slices was used to in the figures and for statistical analysis. The hierarchical clustering was analyzed with Cluster 3.0 ^30^. Heat map was visualized with Java Treeview (version 1.1.6r4) ^31^.

### HCV RNA measurement

HCV RNA was measured with a Food and Drug Administration-approved Abbott Real-Time HCV assay (Abbott Molecular, Des Plaines, IL, USA) as previously described ^20,32^. Fifty ng of total RNA for each sample was input in the assay, and HCV IU/ng total RNA was calculated from the calibration curve of positive controls. The reliable detection limit for HCV with this assay was 1.2 IU/ 50 ng liver total RNA.

### Validation of liver cell types with cell-type-specific genes

Liver cell-type-specific genes for hepatocytes, Kupffer cells (KC), liver sinusoidal endothelial cells (LSEC) and hepatic stellate cells (HSC) were obtained from previous microarray analysis of liver FACS purified cells ^1^ and recent publications of liver single cell RNAseq ^33,34^. Enrichment of the FACS purified liver cell types were confirmed by the selective expression of cell-type-specific genes.

### *Ex vivo* stimulation of liver slices with poly-I:C and LPS

Liver slices were cultured *ex vivo* for 7 days. Final concentrations of 15 μg/mL polyinosinic– polycytidylic acid (poly-I:C) (Sigma, Cat. No. P1530), 1.5 μg/mL lipopolysaccharide endotoxin (LPS) (Sigma, Cat. No. L2630) or 1 × PBS (control, matched by volume) were added to the growth medium. During the stimulation, liver slices were maintained on transwell inserts placed on a rocking platform at 37°C. Three liver slices were harvested at each time point, including 0 h (time zero control), 2 h, 4 h, 8 h, 12 h or 24 h. The concentrations of poly-I:C and LPS treatments were based on our previous studies with isolated liver leukocytes ^35^ and liver slices ^1^.

### Statistical significance test

The non-parametric Mann Whitney test, Kruskal-Wallis test with Dunn’s post test with multiple test comparison correction, and Wilcoxon matched-pairs signed rank test were performed using Prism (version 9.1.0) (GraphPad Software Inc., CA, USA). Multiple comparison correction was not applied in the gene expression analysis, because a specific set of genes were targeted for analysis unlike the standard procedure in a data-driven functional genomics study. In this circumstance, correction for type I error is not advised, because the correction could lead to inflation of type II error (i.e. false negatives) ^36^. The issue of type II errors also arises because of the limited number of tissue samples available to us. For these reasons, we treated each gene we selected as an individual measurement.

## Results

As a first step, we verified the robust response of liver slices during poly-I:C and LPS exposure (**Fig. 1, S2**). The added 2 h time point confirmed that *IFN-α, IFN-β, IFN-γ* and *IFN-λ* was stimulated prior to the elevation of interferon-stimulated genes *IFIT1/2, MX1, CXCL10* and *RSAD2* by poly-I:C. The induction of these antiviral genes was greater with poly-I:C compared with LPS induction (**Fig. 1, S2**). In contrast, *IL1B* and *IL6* were more robustly stimulated by LPS. The response of *IL1B* and *IL6* extended to 24 h post stimulation; in comparison, TNF peaked at 2 h, and gradually declined at 4, 8, 12, 24 h post-stimulation time points (**Fig. S2**). The temporal and gene specificity in poly-I:C and LPS response provided the basis for further dynamic testing of HCV-infected and DAA-cured liver tissue.

**Figure 1.**
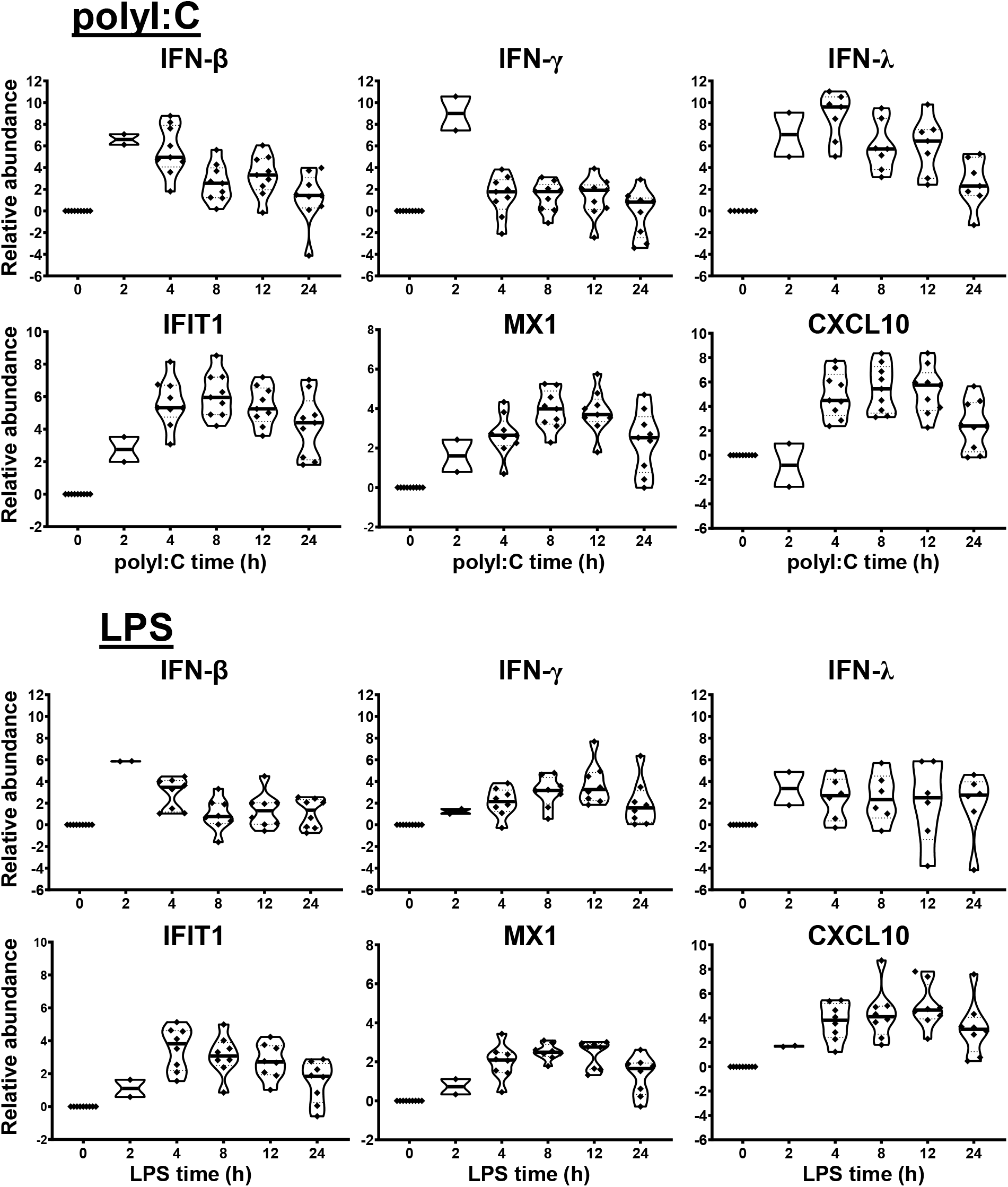
Innate immune response of human liver slices to polyI:C or LPS treatment. Non-HCV-infected specimens are shown. IFN-β, IFN-γ and IFN-λ expression peaked at 2-4 h, whereas interferon-stimulated gene expression for IFIT1, MX1 and CXCL10 peak round 4-12 h. Each marker represents a patient time-point summary which consist of 2-3 biological replicates of liver slices. Relative abundance at the log2 scale is shown, with the median, 25% and 75% quantile values indicated with violin plots.

### Detection and quantitation of HCV RNA with liver slices

Liver tissue from eight chronic HCV-infected patients were studied. All eight patients were diagnosed with primary liver cancer, either hepatocellular carcinoma (HCC) or intrahepatic cholangiocarcinoma (ICC). Four HCV genotypes were identified including HCV G1a, G1b, G2, G6. Likewise, liver tissue from ten DAA-cured patients were studied. The DAA-cured patients completed 8-24 weeks of DAA therapy. All subjects achieve sustained virologic response (SVR) with HCV RNA below the limit of detection after completion of DAA treatment. However, these DAA-cured patients were diagnosed with primary liver cancer (HCC or ICC) post DAA therapy. The time from completion of DAA treatment to liver resection surgery (i.e. the time liver tissue was collected) ranged from less than 1 month to 20 months, with a median of 9 month post-DAA therapy completion (**Table S1**).

To assure the identity of HCV infection, HCV copies and genotype were analyzed by UW virology clinic using the FDA-approved HCV assays. Liver slices originated from the chronic HCV patients were all confirmed with detectable HCV RNA, ranged from 1.2 to 9760.1 with the median value 3325.4 IU/50 ng total RNA. In comparison, none of the non-infected or DAA-treated subjects were positive for HCV RNA *ex vivo* based on the limit of detection of 1.2 IU/50 ng total RNA (**Fig. 2A**). Moreover, chronic HCV-infected liver slices continued to be HCV positive after 7 days of *ex vivo* culture (**Fig. 2B)**, with a range of 1.8-14365.8, and median 405 IU/50 ng total RNA. The viral load on day 7 did not significantly differ from day 0 (Wilcoxon matched-pairs signed rank test, two-tailed, P=0.4688). Previous study showed that HCV can replicate in human liver slices culture for more than 10 days ^37^.

**Figure 2.**
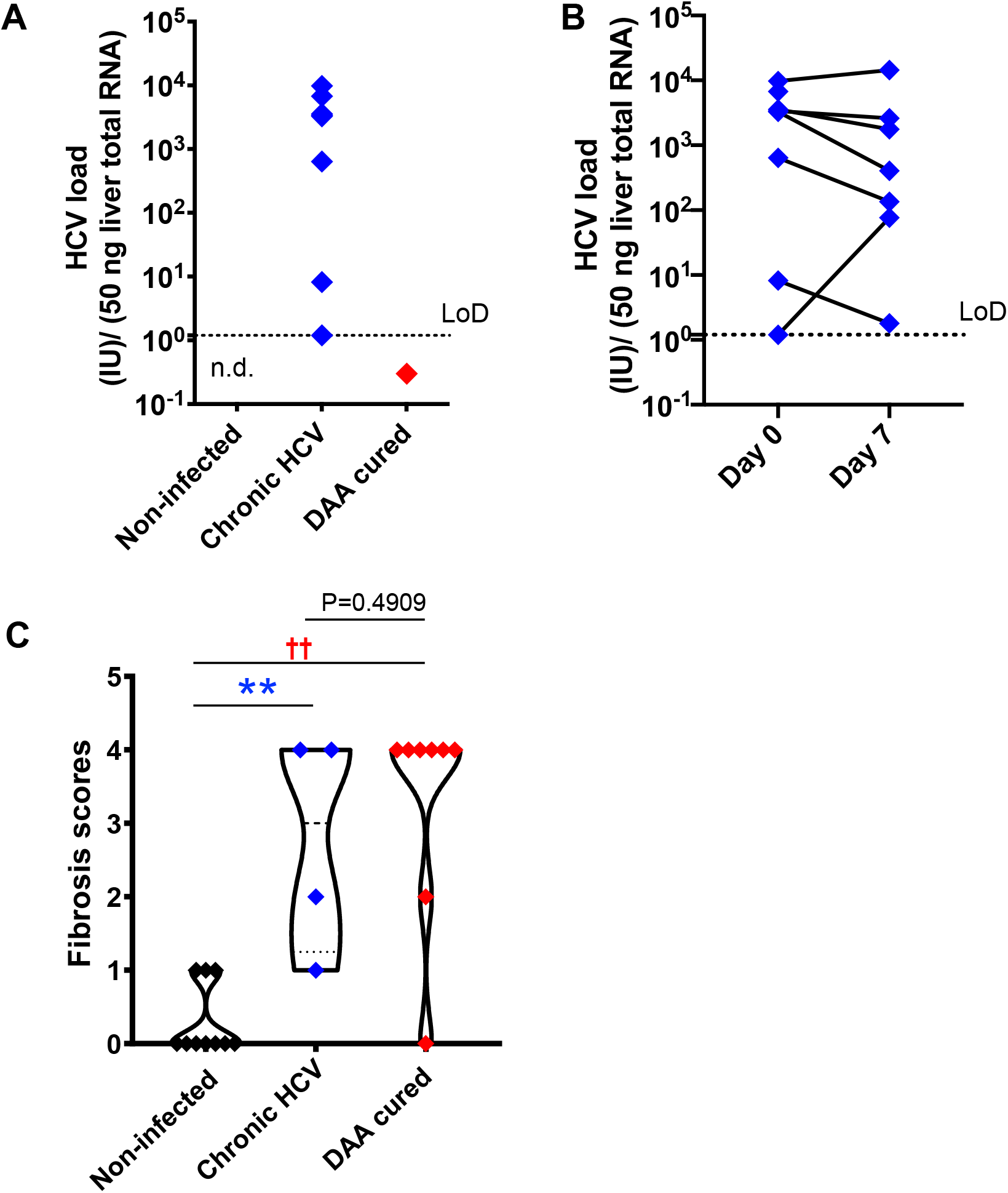
HCV and fibrosis analysis with human liver slices. (A) All eight chronic HCV samples (blue color) were confirmed with positive HCV RNA detection. None of the DAA cured subjects (red color) or the non-infected controls (black color) was positive for HCV RNA. n.d., meaning non-detected. The limit of reliable detection (LoD) was 1.2 IU/ 50 ng liver total RNA. (B) HCV RNA remained robustly detected in the day 7 liver slices cultured from chronically HCV-infected patients. The viral load between day 0 and day 7 liver slices was not statistically significant (P=0.4688, Wilcoxon matched-pairs signed rank test). (C) Fibrosis analysis of the day 0 *ex vivo* liver specimens with trichrome staining and picrosirius red staining indicated that in DAA-treated, and now HCV-negative patients in this study there was persistent fibrosis, similar to untreated HCV and different from tissue without a history of HCV infection. The scoring system was based on the Scheuer/Batts-Ludwig method. Data included 10 non-infected patients, 4 chronic HCV patients, and 8 DAA-treated patients, with the minimum and maximum data points for each subgroup being shown. Statistical significance was based on Mann-Whitney test. **, P < 0.01. ††, P < 0.01

### Fibrosis in chronic HCV-infected and DAA-cured liver tissue

Because chronic HCV infection induced liver cirrhosis, we examined the liver tissue for fibrosis using picrosirius red and Masson’s trichrome staining (**Fig. 2C**). Previous indirect measurements with transient elastography and noninvasive fibrosis indices reported regression of liver fibrosis after DAA treatment ^38,39^. Nevertheless, direct analysis of the liver tissue revealed that six out of eight DAA-cured patients met the definition of cirrhosis (i.e. fibrosis score of 4, Scheuer/Batts-Ludwig method) in livers despite the absence of HCV RNA, even after 20 months after DAA therapy (**Fig. 2C, Fig. 3**). Likewise, the untreated chronic HCV livers were fibrotic as expected ^40^. None of the non-infected subjects had advanced liver fibrosis. Since the duration between the liver tissue collection and the time point of SVR ranged from less than 1 month to 20 months, it remains plausible that the fibrosis will be slowly resolved over a greater time span. As a result of the fibrotic status in HCV-infected and DAA-cured liver tissue, and that HCV-free fibrotic liver tissue was not available in this study, an important caveat is that the abnormal immune gene expression in both the HCV-infected and the DAA-cured liver slices could result either directly from HCV infection, or as a consequence of the fibrosis..

**Figure 3.**
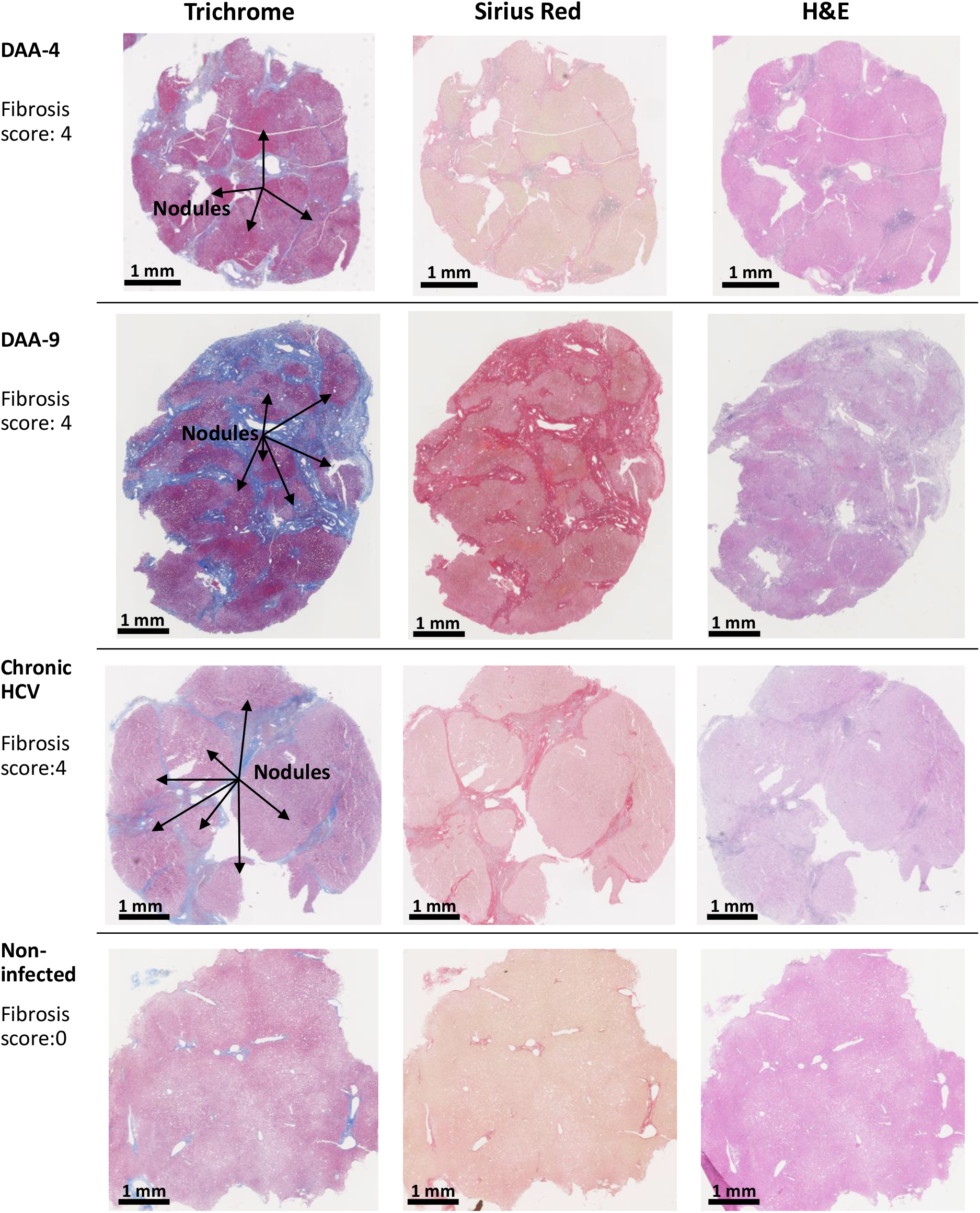
Examples of trichrome stain, picrosirius red stain, and H&E stain analysis of human liver slices. Arrow indicates the nodule formation in the cirrhosis livers. The Scheuer/Batts-Ludwig method was used for fibrosis scoring, with 0: No fibrosis, 1: portal fibrosis, 2: peri-portal fibrosis, 3: bridging fibrosis, and 4: cirrhosis. The DAA-4 liver tissue was collected 12 months after the completion of 24-week DAA therapy, DAA-9 liver tissue was collected 20 months after the completion of 12-week DAA therapy.

### Immune gene abnormality in chronic HCV-infected and DAA-cured *ex vivo*

We used multiplex qRT-PCR to examine 140 genes functioning in immune activation and suppression, and tissue repair. Forty immune genes differed in expression in chronic HCV-infected livers versus non-infected patients (Mann Whitney test, two-tailed, P < 0.05) (**Fig. 4A**). This included antiviral interferon (IFN)-stimulated genes (ISGs) including *IFIT1/2/3, ISG15, RSAD2, MX1, CXCL9, CXCL10*, consistent with previous reports ^41-44^. In addition, immune activation genes including MHC genes *HLA-A, HLA-DRA, CIITA, CD80, CD86*, tumor necrosis factor (TNF) superfamily genes *OX40, 4-1BB, TNFSF9, TNFRSF18* and inflammation genes including *CASP1, IL-12A, IL-12B, TNF, IL-18*, and *NFKB* were also significantly up-regulated in HCV-infected livers (**Fig. 4A**).

**Figure 4.**
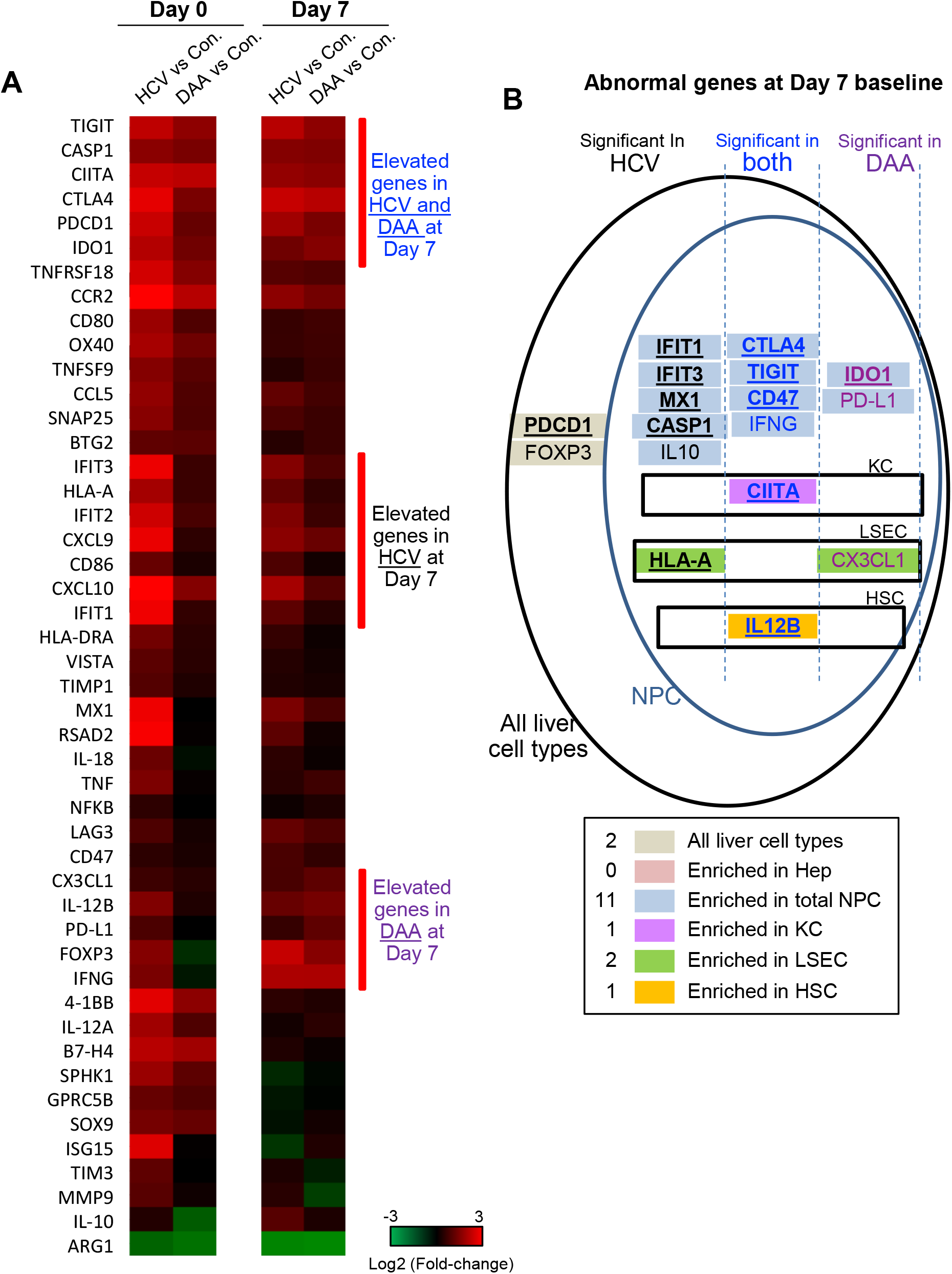
Immune gene changes at the day 7 baseline in chronic HCV-infected and DAA-cured livers. (A) Hierarchical clustering of immune genes with statistically significant elevation in HCV or DAA-cured subjects at Day 0 or Day 7 time points. The group-level fold changes are shown, which were calculated with the mean Ct value of each gene within each patient group. Decreased fold changes are colored in green, and increased fold changes are colored in red. (B) The up-regulated genes in HCV and DAA-cured at day 7 time points. Genes were mapped to liver cell types according to the cell type analysis with day 0 non-infected livers. Genes are grouped by expression patterns in cell compartments and in specimens type. The overlapped genes up-regulated at both day 0 and day 7 time points are bolded and underscored.

DAA treatment restored the antiviral genes to baseline, as reported previously ^45^, but some immune genes were remained elevated after cure of HCV infection. These included immune activation genes (*HLA-A, CIITA, 4-1BB, CCL5, CCR2*), inflammatory genes (*CASP1, IL-12A*), and immune suppression genes (*CTLA4, TIGIT, IDO1, ARG1*). (**Figs. 4A**)

The phosphoinositol-3-kinase (PI3K) pathway-linked genes are susceptible to modification by HCV infection ^18^ and liver fibrosis ^46,47^. We found that *SPHK1, BTG2, SOX9, SNAP25*, and *GPRC5B* were significantly up-regulated in chronic HCV infected livers. Furthermore, *SPHK1, BTG2*, and *SOX9* remained significantly up-regulated after HCV clearance with DAA treatment (**Fig. 4A**).

### Immune gene subsets mapped to hepatic non-parenchymal immune cells

To identify the cellular origin of the dysregulated immune gene subsets, we investigated gene expression with liver specific cells purified from FACS ^1^. Six liver wedge samples of non-infected liver tissue were large enough in size and anatomically ideal for perfusion through the vasculature, which allowed them to be dissociated and analyzed for gene expression. We could not do this with samples of DAA-cured patients due to limitations in the amount of liver tissue obtained. Liver cell types were confirmed with respective cell expression markers^1,33,48^, including *FCN1, NLRP3, ITGA4* for KC, *EMCN, VWF, CD34* for *LSEC, FGFR2, PDGFRB, COL1A1, LRAT* for HSC, and *FBP1, RBP4, ALB* for hepatocytes (**Fig. S3**). Many immune genes that were up-regulated in HCV-infected or DAA-cured fibrotic liver tissue were significantly enriched in the Non-Parenchymal Cell (NPC) compartment (**Table 1, Figs. 4B**). For a smaller subset of immune genes, we were able to map them to specific immune cells with greatest expression, for example *CIITA* mapped to Kupffer cells (KC), *HLA-A* and *CX3CL1* mapped to liver sinusoidal endothelial cells (LSEC), and *IL-12B* mapped to Hepatic Stellate Cells (HSC) (**Fig. 4B, Fig. S4**). Even though NPC may not include the cell types targeted by HCV infection, previous studies demonstrate that NPC can be activated through paracrine signaling from the neighboring HCV-infected hepatocytes ^43,49,50^.

**Table 1.**
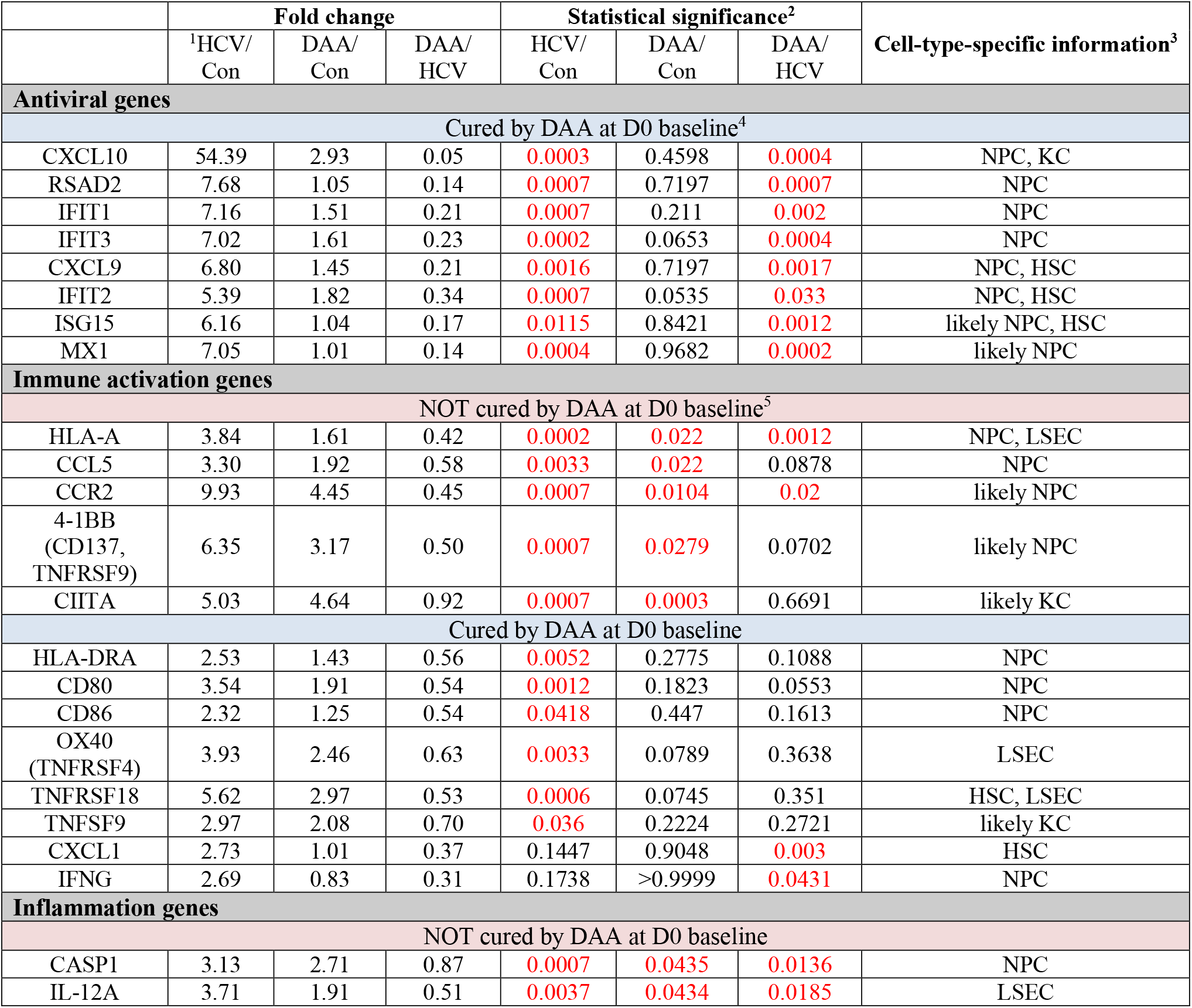
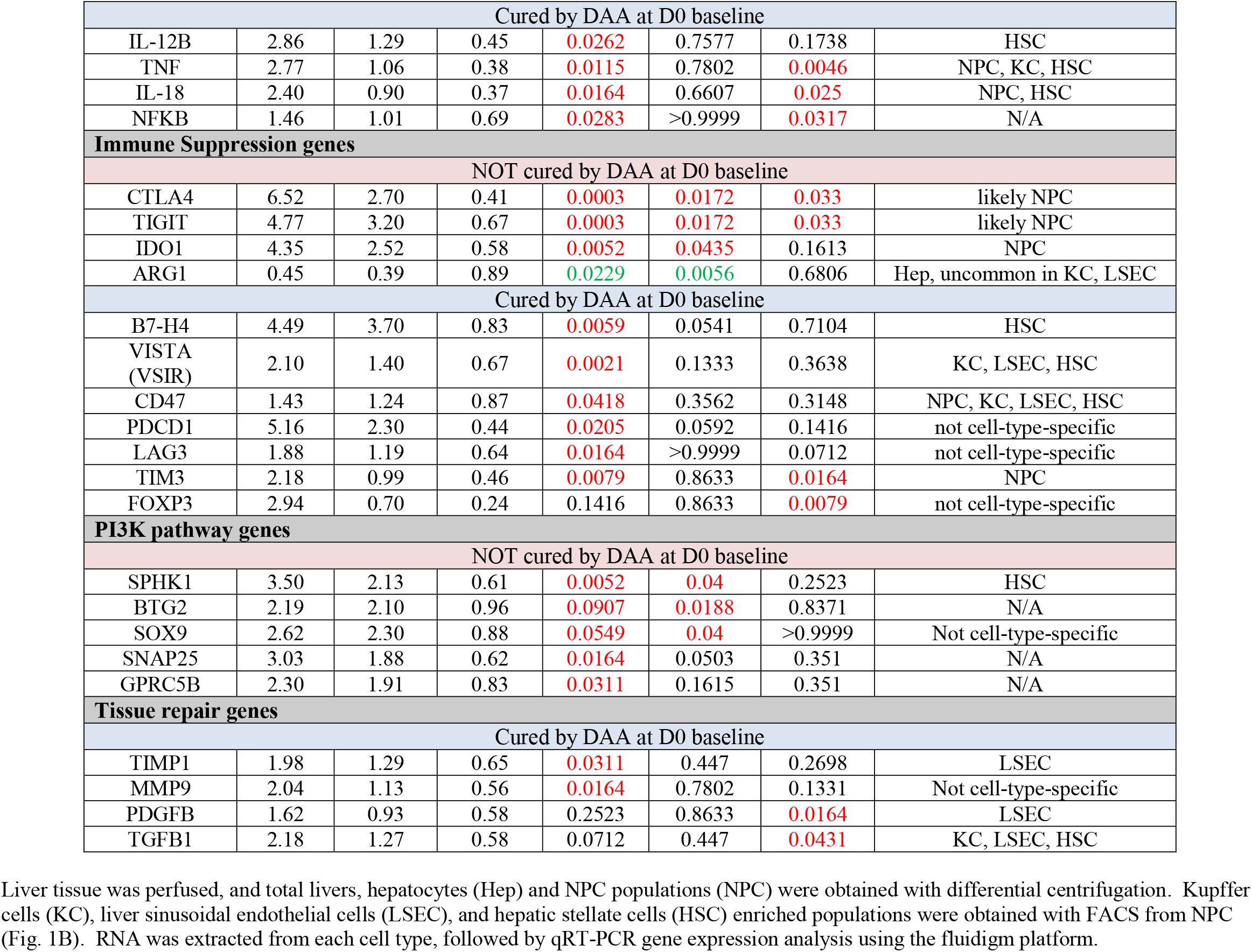
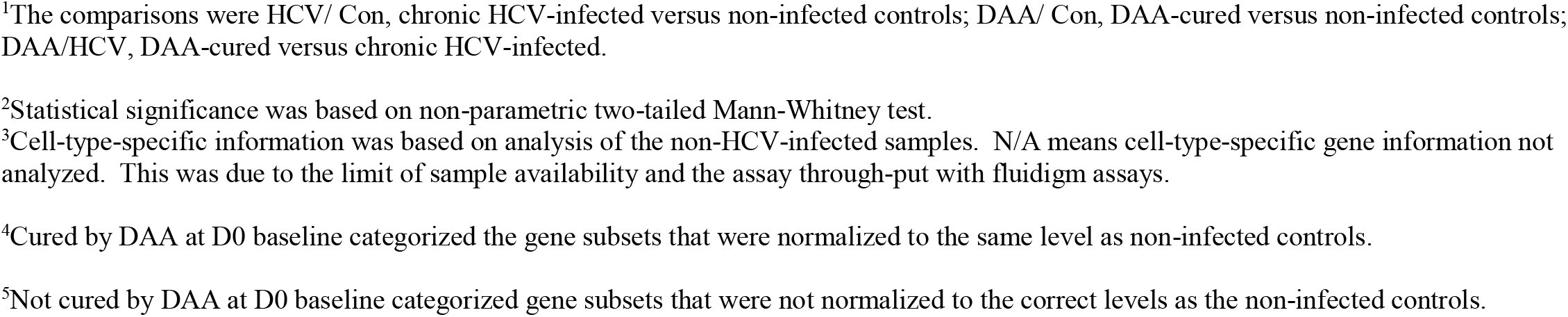
Immune gene changes *ex vivo* (Day 0)

### Innate immune responses were altered in HCV infected and DAA-cured liver tissue

We stimulated liver slices with poly-I:C and LPS for varying times using the method established in our previous study ^1^. We chose day 7 as the time to add stimuli because the immune genes activated by tissue slicing had broadly stabilized after 4 days of *ex vivo* culture ^1^. The immune genes were recorded at the day 7 baseline prior to the treatment. A number of the immune genes remained at elevated baseline in HCV-infected or DAA-cured samples compared with non-infected control (**Fig. 4AB**). The PBS mock stimulation did not significantly induce IFNs and ISGs in liver slices ^1^. Notably, expression of TLR3, TLR4 themselves and NF-κB transcription factor were not significantly different at day 7 among chronic HCV-infected, DAA-treated and non-infected liver slices (**Fig. S5A**). We used the ΔCt method with internal normalization to *ACTB, GAPDH* and *HPRT* to evaluate the overall abundance level post stimulation (**Fig. S6**). We also used the ΔΔCt method against the day 7 T_0_ to assess the enhanced net increase of genes during TLR3/TLR4 response (**Fig. 5**). We confirmed that TLR3 and TLR4 are primarily expressed in the NPC compartment (**Fig. S5B**).

**Figure 5.**
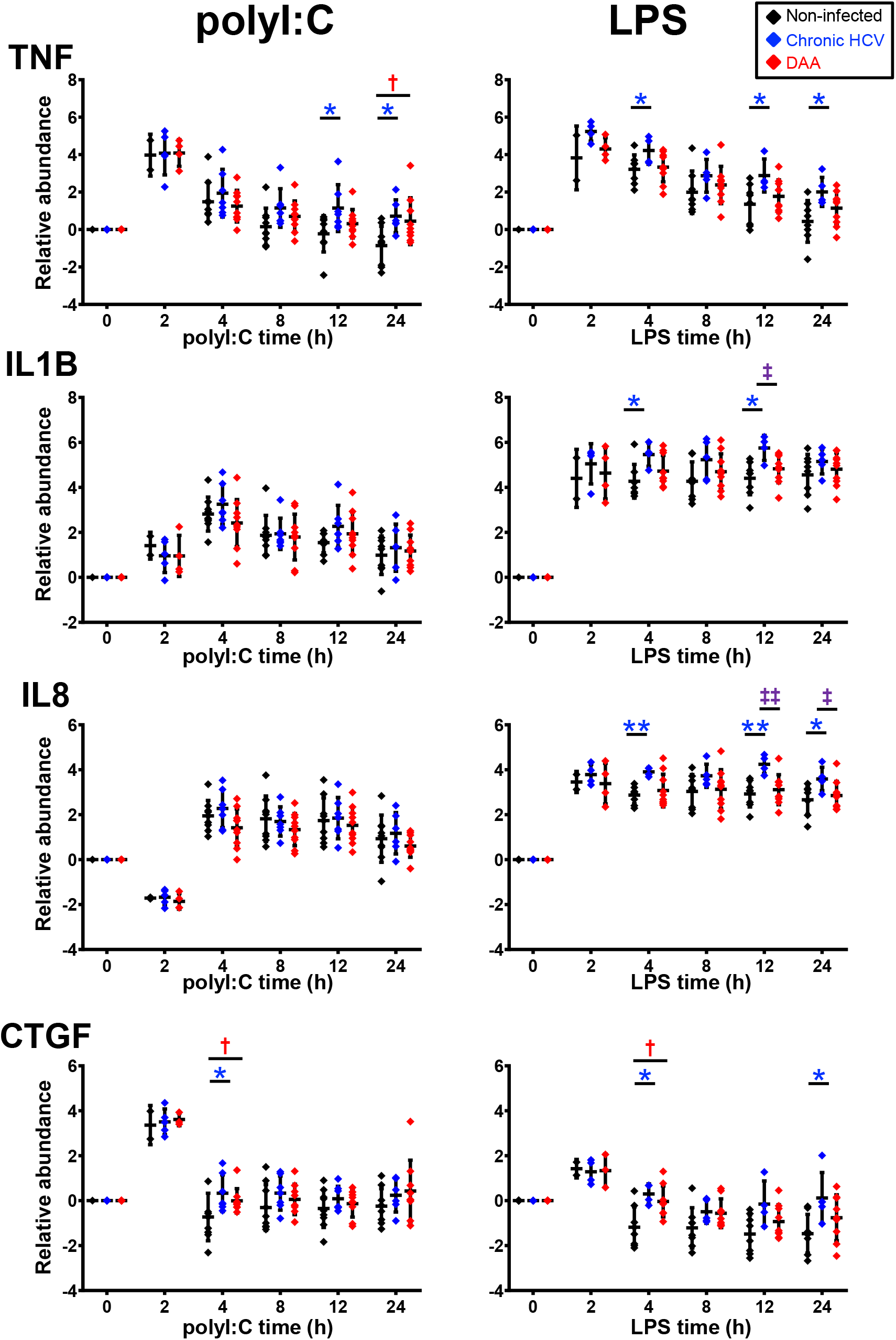
Delta-delta Ct analysis showing up-regulated genes post-poly-I:C or LPS stimulation in chronic HCV-infected and DAA-cured liver slices. The relative gene abundance was normalized to to the arithmetic mean of Ct values of ACTB, GAPDH and HPRT1. The relative abundance was further normalized to the day 7 time zero measurements. Statistical significance was based on two-tailed Mann-Whitney test. *, statistical significantly different between non-infected versus chronic HCV liver slices. †, statistical significantly different between non-infected versus DAA-cured liver slices. ‡, statistical significantly different between chronic HCV versus DAA-cured liver slices. Levels of statistical significance *, P <0.05 **, P<0.01, ***, P<0.001. The mean and standard deviation within each group are plotted.

First, robust induction of IFNs and ISGs by both TLR3 and TLR4 agonists was observed in liver slices from all three groups of liver specimens (*i*.*e*., non-infected, HCV-infected, and DAA-cured). The sequential signaling cascade induced by TLR3 and TLR4 pathways described in **Fig. 1** and **Fig. S1** for non-infected liver tissue, were also pronounced in the HCV-infected and DAA-cured liver tissue. For example, *IFNB1* and *IFNL3* (IL28B) peaked at 2-4 h, prior to the maximum abundance of ISGs (*IFIT 1/2/3, RSAD2* and *MX1*) which occurred at 4-8 (**Figs. 5 and S6**).

Second, some immune genes were not suppressed but induced more strongly by TLR3 agonist in liver slices of chronic HCV-infected liver at least at one time point (Mann-Whitney test, two tailed, P < 0.05). These included antiviral genes *MX1, RSAD2, IFNG*, chemokine genes *CCL3, CX3CL1, CXCL9*, inflammatory genes *TNF, CASP1* and *IL-10* (**Fig. S7**). Other common activation markers for TLR3 signaling, including *IFNB1, IL28B, IFIT 1/2/3, ISG15, CXCL10, IL-1B* and *CCL-5*, were also responsive in chronic HCV-infected slices with elevation of greater than 10-fold, as strong as changes in the non-infected liver slices. Thus, TLR3 sensing and signaling did not appear to be impaired in chronic HCV-infected liver slices. Furthermore, *TNF, IL-10* and *RSAD2* remained abnormally induced at 12 h and 24 h in chronic HCV liver slices, indicating persistent inflammation.

TLR4 sensing and signaling were similarly robust in chronic HCV-infected livers. *IFIT1, IFIT3, RSAD2, MX1, CCL3, CASP1* and *IL-10* were again identified along with *IL-8, IL-1B, IL-1A* and *IL12B* at greater abundance post LPS stimulation in chronic HCV-infected liver slices on at least one time point (Mann-Whitney test, two tailed, P < 0.05) (**Fig 6A**).

**Figure 6.**
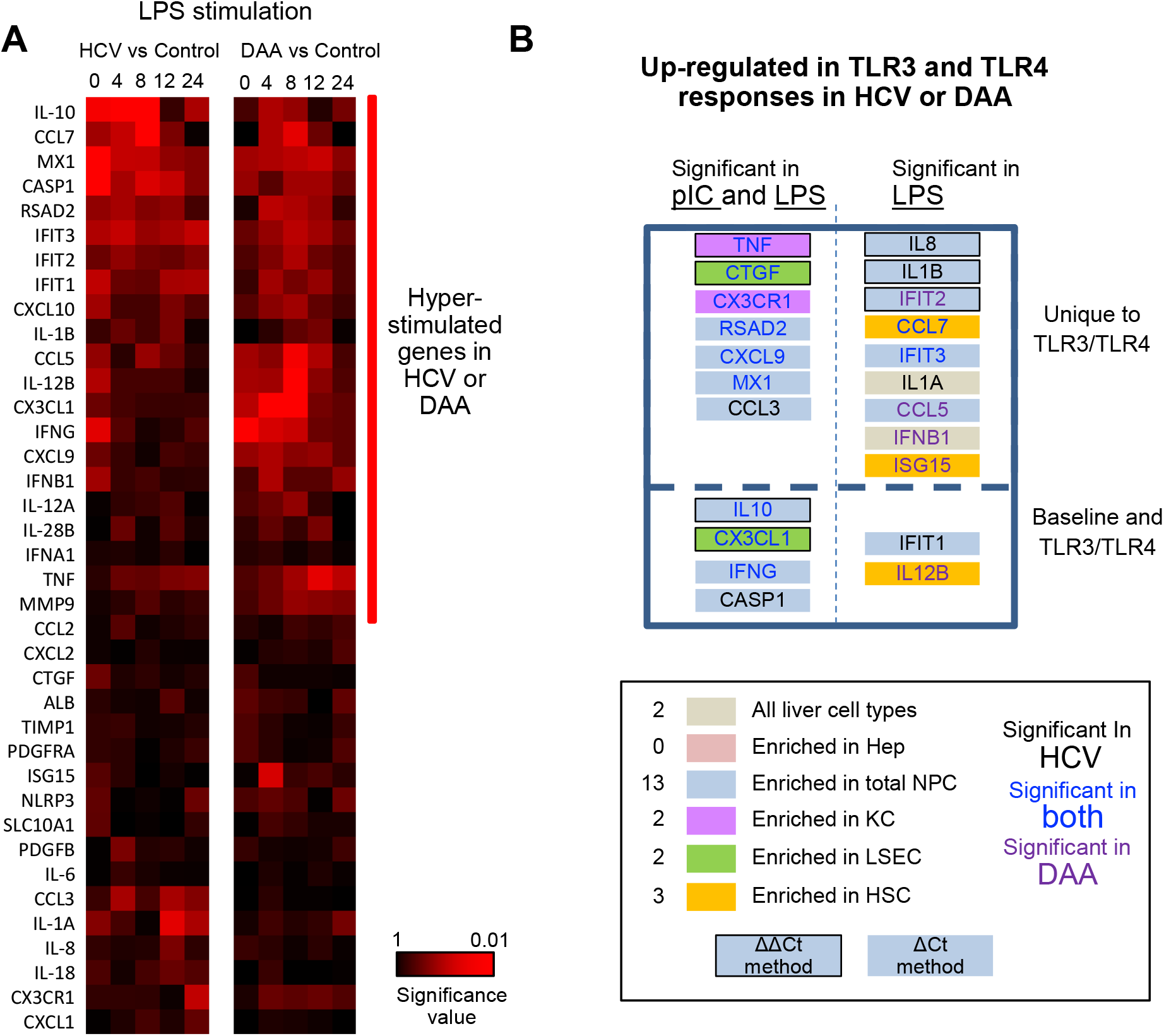
Altered TLR3 and TLR4 response in chronic HCV-infected liver slices not restored in DAA-cured liver slices. (A) Hierarchical clustering of immune responsive genes with 2-fold or greater induction during LPS stimulation. The statistical significance P value with ΔCt method is shown. Gene abundance of individual liver slices was normalized to the arithmetic mean of Ct values of ACTB, GAPDH and HPRT. Two-tailed Mann-Whitney test was used. Greater significance P value is colored with darker red. (B) Immune genes up-regulated in polyI:C or LPS treatment were mapped to live cell compartment according to the day 0 information. Genes identified with ΔΔCt method, i.e. with an enhanced net increase during TLR3/TLR4 response is highlighted with a black bordering. Genes elevated in HCV- or DAA-samples or both are colored accordingly. Elevated genes were grouped by whether they were differentially expressed in TLR3 or TLR4 treatment not at the baseline level, and whether polyI:C or LPS treatment revealed differential responses. Antiviral genes RSAD2, CXCL9, MX1, IFIT3 were corrected at baseline in DAA-cured samples, but manifested abnormal up-regulation during polyI:C and LPS treatment.

Third, most of the hyper-induction of the genes in HCV-infected tissue (9/11 genes in TLR3 response, 12/18 genes in TLR4 response) was not reversed in the tissue from formerly HCV+, but now successfully treated donors. *IFIT2, IFNB1, CCL5, ISG15* and *IL12B* were more strongly induced by LPS in DAA-cured tissue, previously not identified with HCV-infected tissue (**Fig. 6B**).

In terms of the ΔΔCt net increase, *TNF, CTGF, IL-10, CX3CL1* were more strongly induced by both polyI:C and LPS in both HCV-infected and DAA-cured liver tissue. *IL-1B* and *IL-8* in HCV-infected, and *IFIT2* in DAA-cured liver tissue were more responsive to LPS (**Fig. 6B**). Importantly, a subset of these genes *TNF, CX3CR1* in KC, *CTGF* in LSEC, *CCL7, ISG15* for HSC, and *IL-8, IL-1B, RSAD2, CXCL9, CCL3* for NPCs were only revealed through the TLR3 and TLR4 stimulation assays, but not with the *ex vivo* day 0 measurements. These results highlighted the unique value to measure dynamic changes with liver tissue to reveal immune signaling defects (**Fig. 6B**).

## Discussion

In this study, we pioneered the PCLS technology to study innate immune response with the normal and pathological liver tissue. We showed that immune abnormalities persist after the elimination of HCV by anti-viral therapy, and the persisting inflammatory and immune signatures mapped to hepatic non-parenchymal cells. Innate immunity was not suppressed but enhanced in HCV-infected tissue, and these abnormalities were not corrected after effective DAA treatment.

In human hepatocyte cultures ^51,52^ HCV infection triggered cleavage of MAVS and TRIF host proteins by HCV NS3/4A protease, disrupting TLR3-antiviral signaling. Reduction of protein levels of MAVS and TRIF was also observed previously in chronic HCV-infected livers ^53,54^. Nevertheless, we observed a robust response to TLR3 signaling in the HCV-infected liver, which supports the idea that ISG induction is weak in HCV infected cells, but strong in HCV infected liver due to paracrine activation of NPC. The data on the direct effect of HCV on the TLR3 response in liver tissue are novel and distinct from previous work focused on the IFN-induced gene expression in chronically HCV-infected liver by pegylated IFN therapy ^55,56^, since such direct activation of IFN signaling bypasses MAVS and TRIF.

A previous study reported that HBV does not interfere with innate immune response based on chronic HBV-infected liver biopsy tissues ^57^. However, in chronic HCV-infected tissue, we found innate immunity to TLR3 and TLR4 was not suppressed but enhanced. The different outcome may reflect the differences in immune evasion mechanisms between DNA and RNA viruses. Furthermore, the tissue status and treatment method were not identical between two studies. In chronic HCV-infected tissue, advanced fibrosis was observed, and this study also used the day 7 as treatment time zero when immune activation by tissue slicing was broadly stabilized.

From the days of IFN-based therapy, we know that HCV-induced liver disease can regress when the virus is eradicated from the liver ^58^. However, halting or reversing liver disease upon IFN cure of HCV has not been universal; some patients with advanced disease still go on to develop HCC ^59-61^. DAA drugs are now able to cure the majority of HCV infections, even in subjects with advanced liver disease. Nevertheless, clearance of HCV with DAA treatment may only partially restore immune cell function. In this study, we have identified immune genes that were restored to normal by DAA treatment, and more importantly immune genes that were not restored *ex vivo* (*i*.*e*., at day 0 time point), including a subset of immune activation genes, inflammation genes, immune suppression, and PI3K pathway genes.

The era during which some, but not all HCV patients were treated with DAA was brief and has passed. Within this narrow time window, we worked with tissue from patients who were treated for surgical resection of a liver lesion. Accordingly, there are inherent limitations to this study. First, due to the small sample size, we did not further classify patient subgroups based on HCV genotypes. Likewise, other subject characteristics including gender, age, genetic polymorphisms (such as *IL28B*), and liver fibrosis status, all of which may affect immunity and responses to HCV infection ^14,62,63^ were not used as factors to subdivide patient groups in the downstream analysis. Surprisingly, we observed significant effects that cut across these hidden variables, with consistent identification of aberrant immune gene at baseline day 0 and day 7, as well as those with altered TLR3 and TLR4 responses. Second, we cannot determine how far the sustained abnormality is due to the host response to hepatic fibrosis or HCC rather than HCV-mediated. The DAA-cured subjects we studied were based only on those who developed HCC or ICC post DAA therapy, since these were the patients who were subject to liver resection. Thus, our DAA-cured samples only covered a unique sub-group of DAA-cured patients. Future study may extend the conclusion through analysis of a more diverse group of clinical situations, including chronic HCV patients and DAAs-SVR cases with or without fibrosis or HCC, as well as non-HCV related liver tissues with fibrosis or HCC. Liver tissue with longer time span post DAA therapy (>20 months) will also help clarify the resolution of liver cirrhosis by DAA therapy.

We made use of the PCLS approach opportunistically to obtain data from the group of untreated HCV-infected subjects, a group that has now disappeared from the surgical patient pool. While many of the scientific conclusions need to be tentative because of the heterogeneity of both subjects and controls, this study demonstrates the power of PCLS to document functional responses of intact human liver tissue. With appropriate selection of tissue sources, it will add much to the study of human liver immunopathology.

## Data Availability

All relevant data are within the manuscript and its Supporting Information files.

## Acknowledgements

We thank Pathology Research Services Laboratory, and Histology and Imaging Core (HIC) at UW for their help in data and image acquisition.

## Authors’ contributions

X.W., J.B.R., A.C., H.H., R.S.Y. and I.N.C. designed research; X.W., J.B.R., A.K., A.L.G., N.H., H.L.K, M.T. and A.C. performed research; K.R.J., S.J.P. and R.S.Y. contributed reagents/analytic tools; X.W., A.L.G., C.D.T. and I.N.C. analyzed data; X.W., S.J.P. and I.N.C. wrote the manuscript; all authors reviewed the manuscript and provided suggestions.

## Abbreviations

Abbreviations are provided for some terms not commonly known; some are also explained in the manuscript. Authors will conform with journal standards if paper is accepted scientifically.

PCLS: precision-cut liver slice
TLR: toll-like receptor
NPC: non-parenchymal cell
KC: Kupffer cells
LSEC: Liver sinusoidal endothelial cells
HSC: hepatic stellate cells
HCV: hepatitis C virus
DAA: direct-acting antiviral
ISGs: interferon-stimulated genes
HCC: hepatocellular carcinoma
ICC: intrahepatic cholangiocarcinoma
SVR: sustained virologic response

