## Supplemental Figures 1-7 for "Response of human liver tissue to innate immune stimuli"

Figure S1

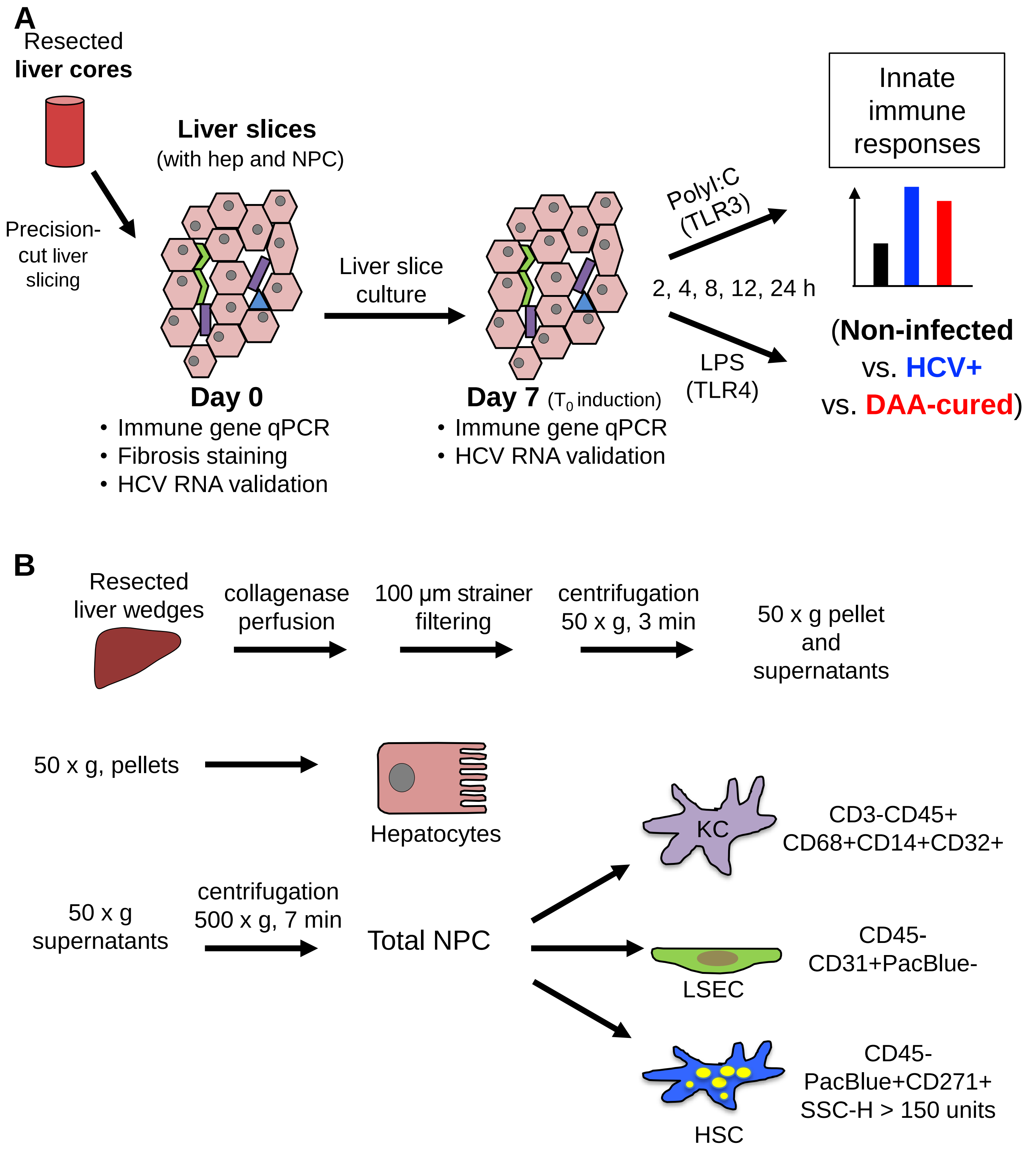


**Figure S1. Experimental Overview for this study**. (A) Human liver slices to study innate immune response. Liver slices were harvested on Day 0 immediately after slicing, or cultured for seven days until ex vivo stimulation with TLR3 agonist poly-I:C or TLR4 ligand LPS. Innate immune responses were compared among three groups of tissue samples including non-infected patients (controls), chronic HCV-infected patients (HCV+), and patients with previous history of HCV infection who were cured by DAA treatment (DAA-cured patients). (B) Specific liver cell types were purified from fresh resected liver wedges with perfusion, differential centrifugation and fluorescence-activated cell sorting (FACS) techniques. Immune genes of interest were analyzed with liver cell samples to determine liver cell types with enriched gene expression. Hepatocytes (Hep), total non-parenchymal cells (NPC), Kupffer cells (KC), liver sinusoidal endothelial cells (LSEC), or hepatic stellate cells (HSC).

Figure S2

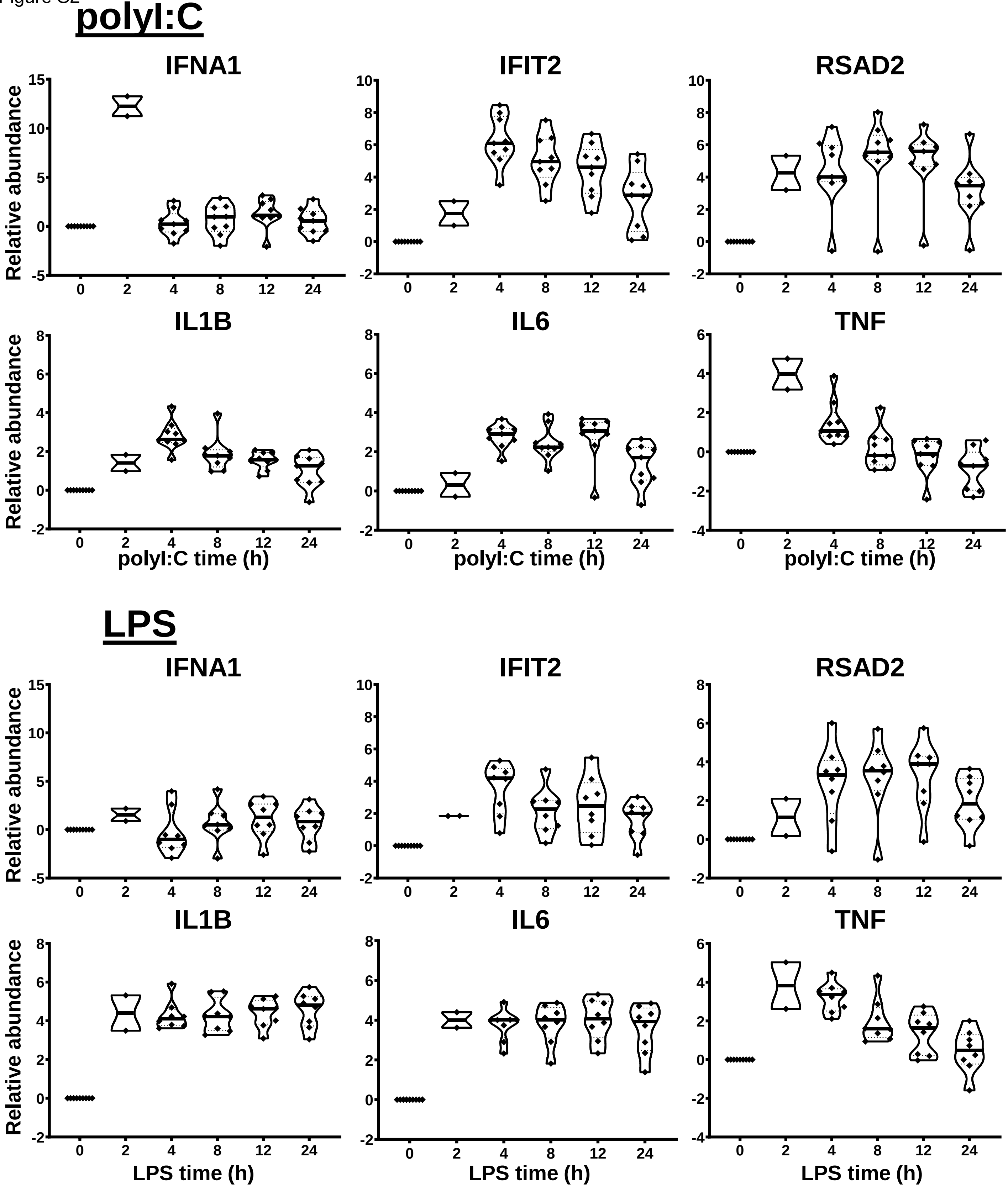


**Figure S2. Additional examples for innate immune response of human liver slices to polyI:C or LPS treatment.** Non-HCV-infected specimens are shown. IFNA1 peaked at 2 h, while IFIT2 and RSAD2 peaked at 4-12 h. IL1B, IL6 and TNF expressions were more robustly stimulated with LPS treatment compared with polyI:C, with a peak induction 2 h onwards. Each marker represents a patient time-point summary. Relative abundance at the log2 scale is shown, with the median, 25% and 75% quantile values indicated with violin plots.

Figure S3

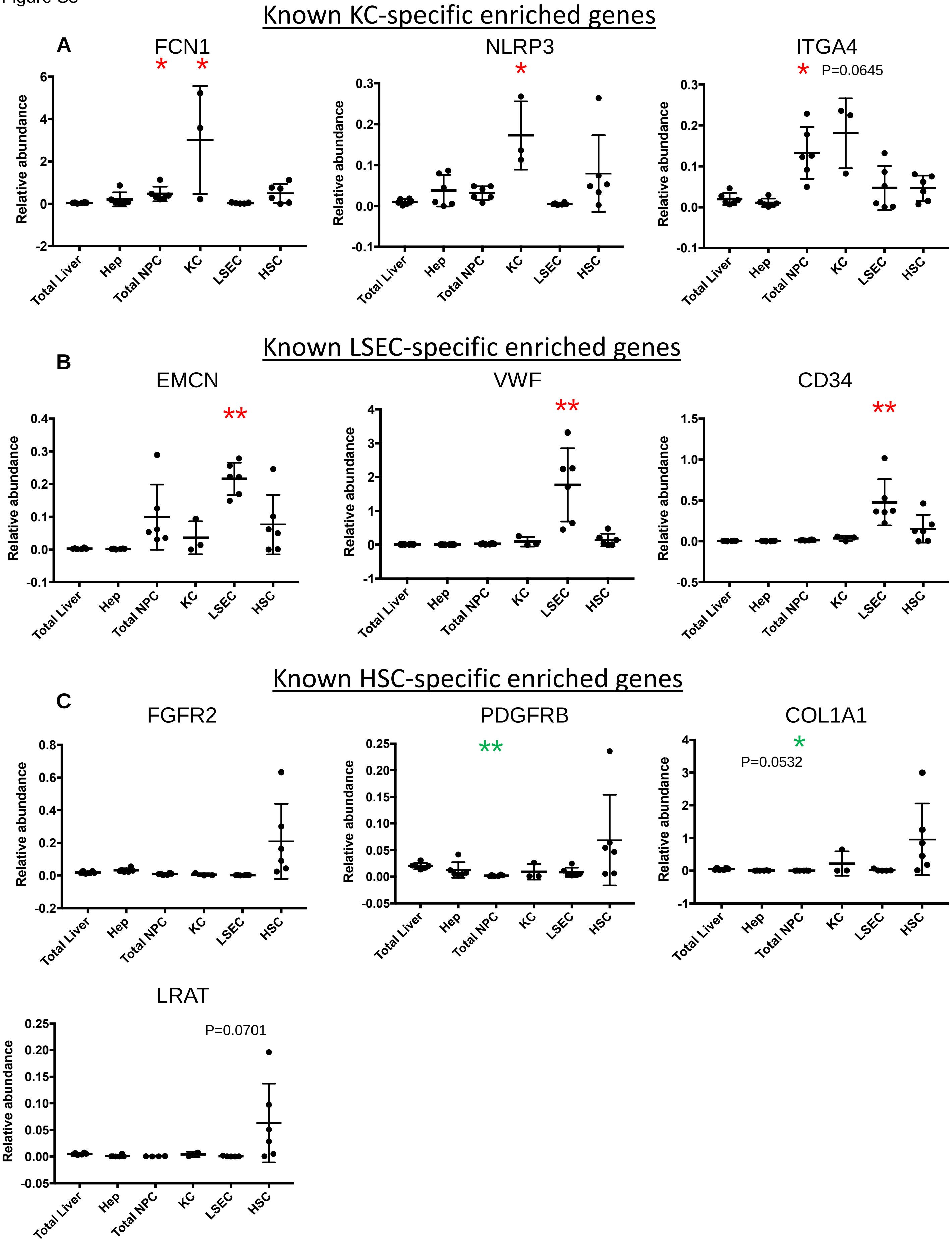
Known KC-specific enriched genes
A
FCN1
NLRP3
ITGA4
P=0.0645
Relative abundance
Total Liver
Hep
Total NPC
KC
LSEC
HSC
Known LSEC-specific enriched genes
B
EMCN
VWF
CD34
Known HSC-specific enriched genes
C
FGFR2
PDGFRB
COL1A1
P=0.0532
LRAT
P=0.0701

Figure S3

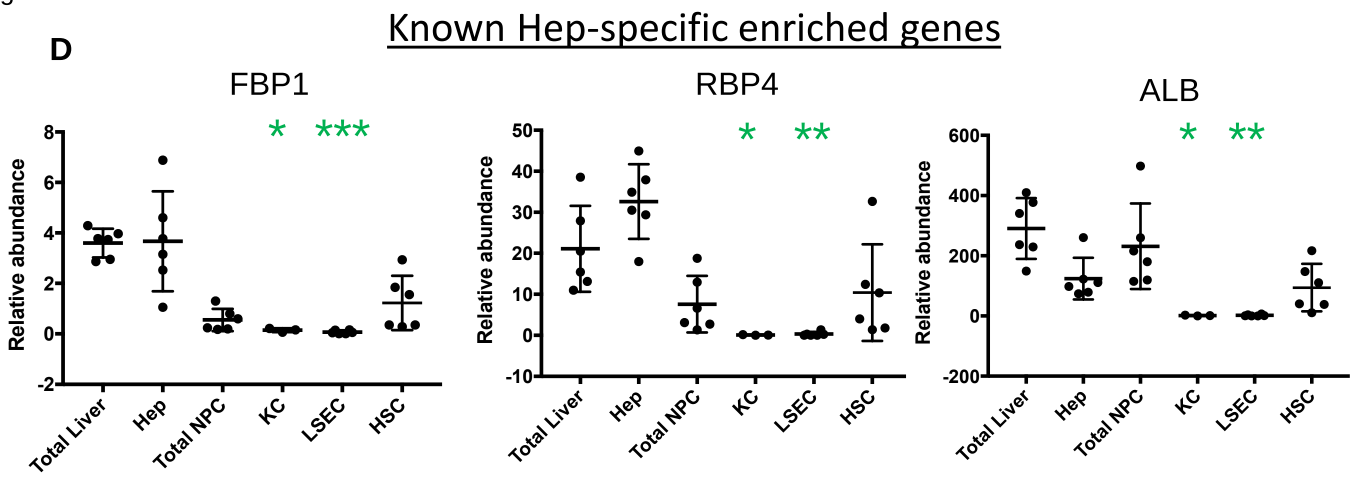


**Figure S3. Validation of FACS purified liver cell types with cell-type-specific gene markers.** The compared samples included total liver samples, hepatocytes (Hep), total non-parenchymal cells (NPC), Kuffer cells (KC), liver sinusoidal endothelial cells (LSEC), and hepatic stellate cells (HSC). See Methods for purification details. Each dot within each cell type represents an individual patient sample. The relative gene abundance was normalized to arithmetic mean of ACTB, GAPDH and HPRT1. The relative abundance of linear scale is shown. Statistical significance was based on Kruskal-Wallis test with Dunn's post test with multiple test comparison correction. Genes that were enriched in a cell type compared with total livers were denoted with red asterisks, while genes that were significantly less abundant compared with total liver were indicated with green asterisks. *, P <0.05 **, P<0.01, ***, P<0.001. The mean and standard deviation were plotted. Graphs and statistical test were analyzed with Prism version 9.1.0. Albumin which was thought as a good hepatocyte specific marker were detected with positive signals in cell fractions of total NPC and HSC. Thus, ALB is less hepatocyte-specific compared with FBP1 and RBP4.

Figure S4

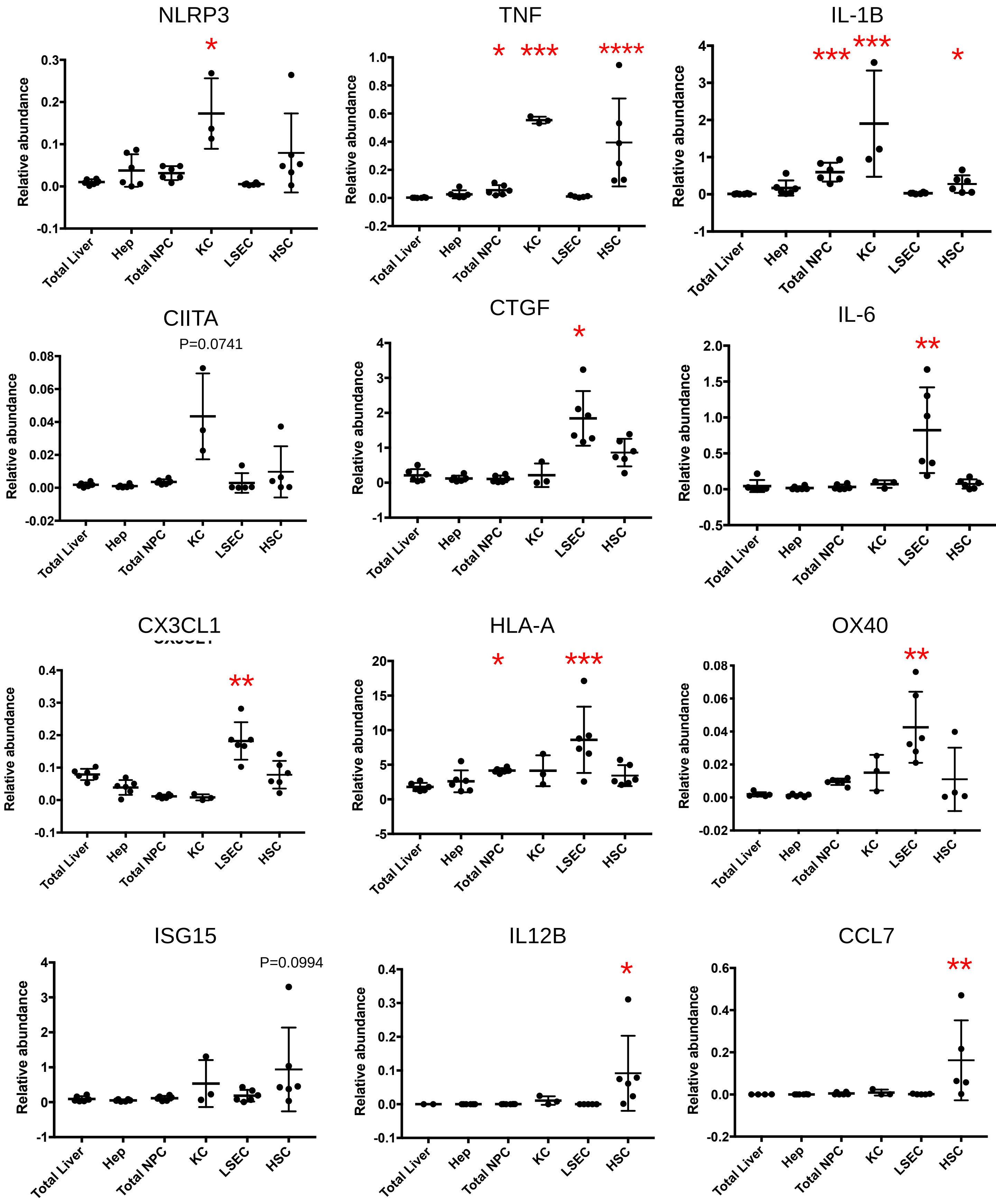
NLRP3
*
Relative abundance
0.3
0.2
0.1
0.0
-0.1
Total Liver
Hep
Total NPC
KC
LSEC
HSC
TNF
*
***
****
Relative abundance
1.0
0.8
0.6
0.4
0.2
0.0
-0.2
Total Liver
Hep
Total NPC
KC
LSEC
HSC
IL-1B
***
***
*
Relative abundance
4
3
2
1
0
-1
Total Liver
Hep
Total NPC
KC
LSEC
HSC
CIITA
P=0.0741
Relative abundance
0.08
0.06
0.04
0.02
0.00
-0.02
Total Liver
Hep
Total NPC
KC
LSEC
HSC
CTGF
*
Relative abundance
4
3
2
1
0
-1
Total Liver
Hep
Total NPC
KC
LSEC
HSC
IL-6
**
Relative abundance
2.0
1.5
1.0
0.5
0.0
-0.5
Total Liver
Hep
Total NPC
KC
LSEC
HSC
CX3CL1
**
Relative abundance
0.4
0.3
0.2
0.1
0.0
-0.1
Total Liver
Hep
Total NPC
KC
LSEC
HSC
HLA-A
*
***
Relative abundance
20
15
10
5
0
-5
Total Liver
Hep
Total NPC
KC
LSEC
HSC
OX40
**
Relative abundance
0.08
0.06
0.04
0.02
0.00
-0.02
Total Liver
Hep
Total NPC
KC
LSEC
HSC
ISG15
P=0.0994
Relative abundance
4
3
2
1
0
-1
Total Liver
Hep
Total NPC
KC
LSEC
HSC
IL12B
*
Relative abundance
0.4
0.3
0.2
0.1
0.0
-0.1
Total Liver
Hep
Total NPC
KC
LSEC
HSC
CCL7
**
Relative abundance
0.6
0.4
0.2
0.0
-0.2
Total Liver
Hep
Total NPC
KC
LSEC
HSC

Figure S4

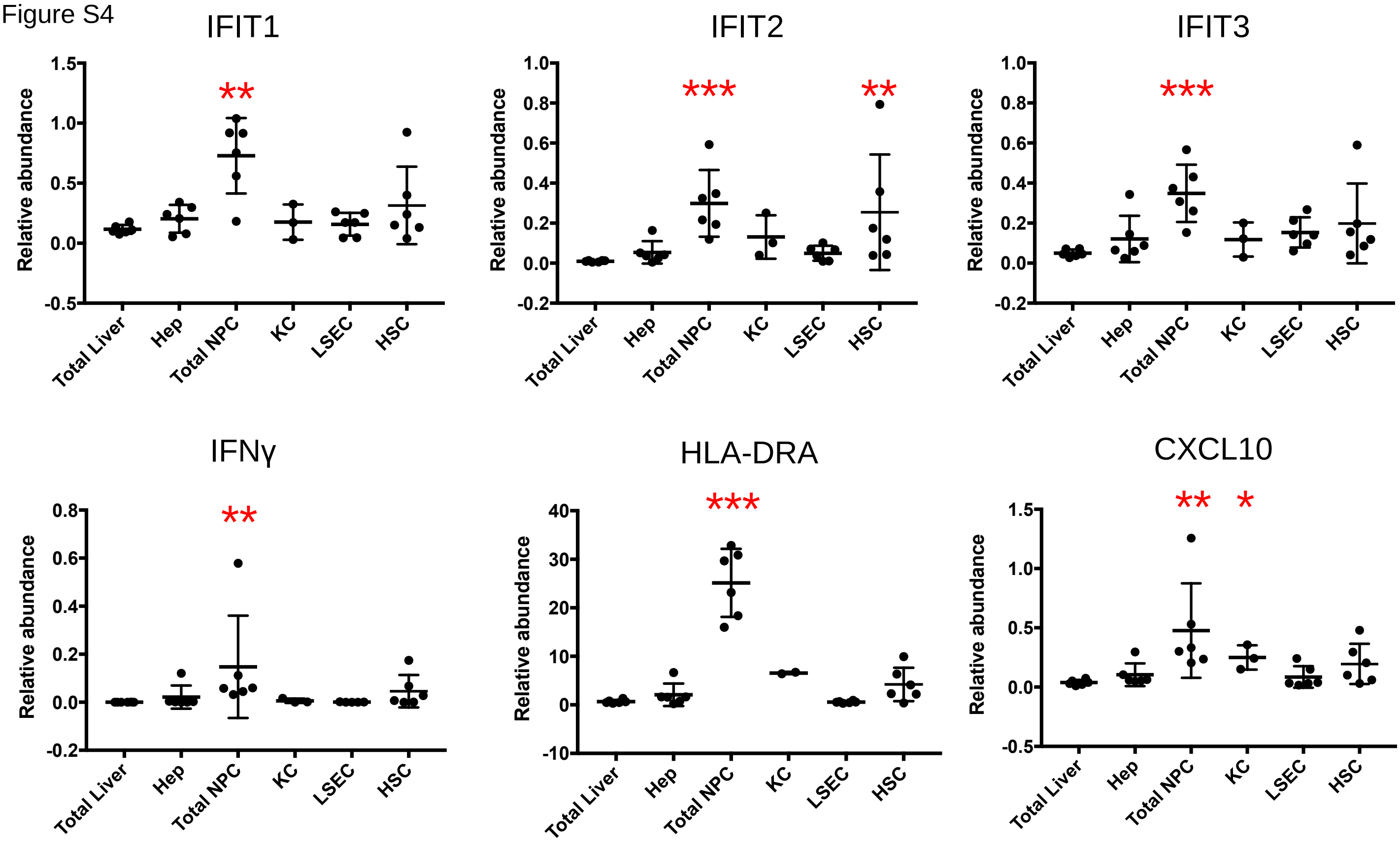


**Figure S4. Gene expression detected with FACS purified liver cells.** Hepatocytes (Hep), total non-parenchymal cells (NPC), Kuffer cells (KC), liver sinusoidal endothelial cells (LSEC), and hepatic stellate cells (HSC). Each dot within each cell type represents a biological replicate. The relative gene abundance was based on the delta delta Ct of the arithmetic mean of ACTB, GAPDH and HPRT1. Statistical significance was based on Kruskal-Wallis test with Dunn's post test with multiple test comparison correction, performed in Prism 9.1.0 . Genes that were enriched in a cell type compared with total livers were denoted with red asterisks *, P <0.05 **, P<0.01, ***, P<0.001. The mean and standard deviation were plotted.

Figure S5

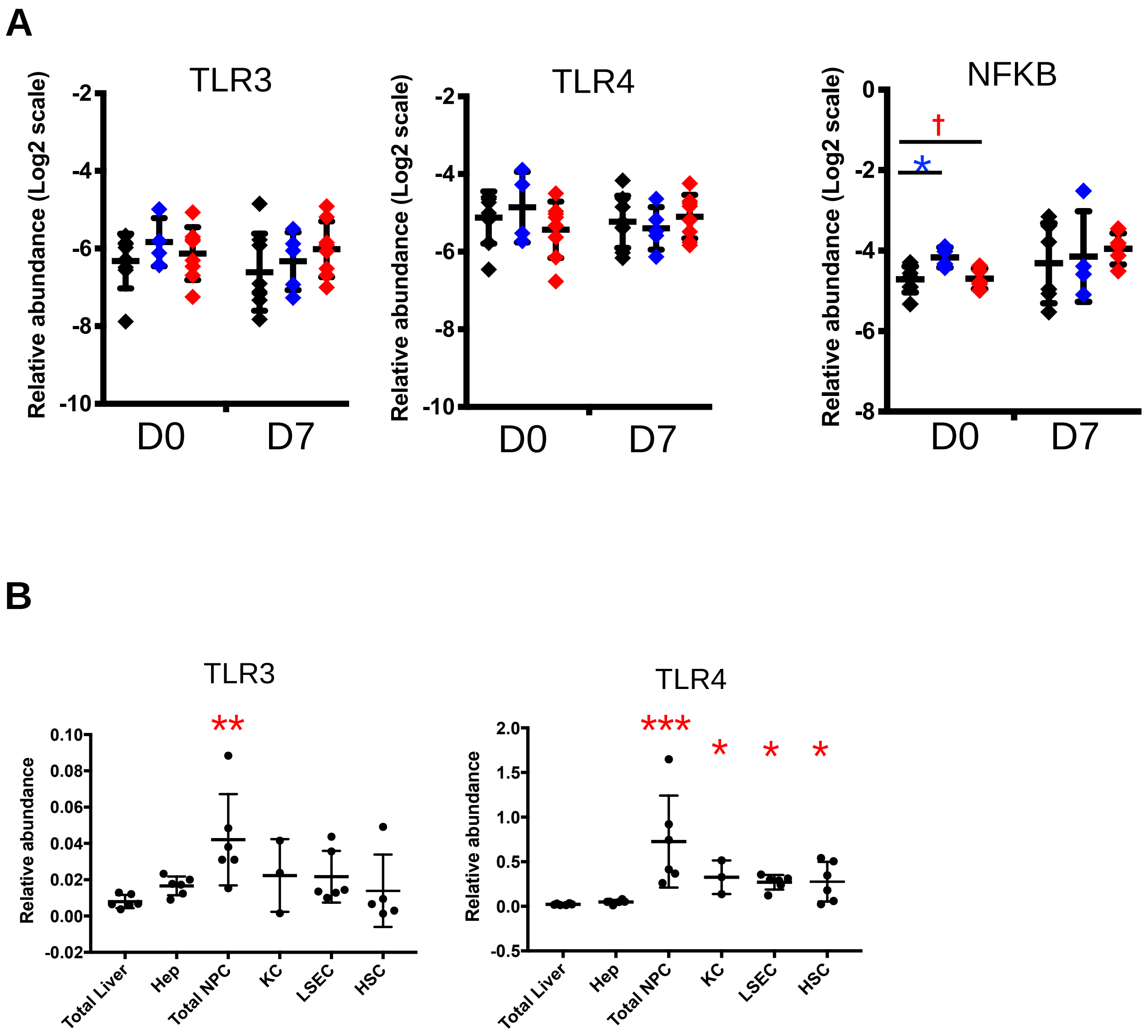


**Figure S5 TLR3 LTR4 in liver slices.** (A) TLR3, TLR4 no change at baseline D0 and D7. NFKB differed at Day0; seven days cultured normalized the differences. The relative abundance is showed at log2 scale. The mean and standard deviation are shown. (B) TLR3, TLR4 expressed at NPC compartment.

Figure S6

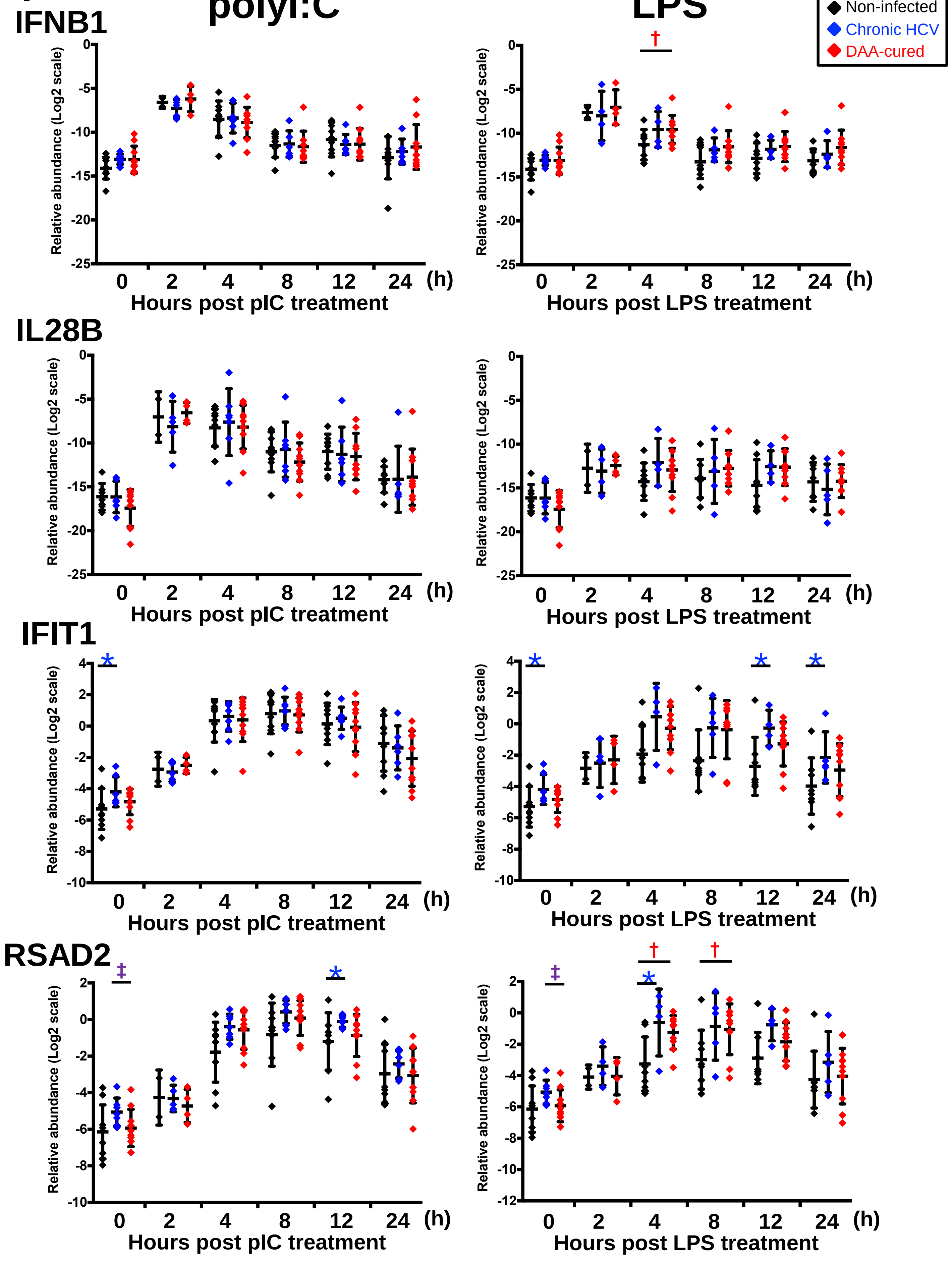

polyI:C
LPS
Non-infected
Chronic HCV
DAA-cured
IFNB1
IL28B
IFIT1
RSAD2
Relative abundance (Log2 scale)
Hours post pIC treatment
Hours post LPS treatment
0
2
4
8
12
24
(h)
*
†
‡

**Figure S6. Delta-Ct analysis showing up-regulated genes post- poly-I:C or LPS stimulation in chronic HCV-infected and DAA-cured liver slices.** The relative gene abundance was normalized to to the arithmetic mean of Ct values of ACTB, GAPDH and HPRT1. Statistical significance was based on two-tailed Mann-Whitney test. *, statistical significantly different between non-infected versus chronic HCV liver slices. †, statistical significantly different between non-infected versus DAA-cured liver slices. ‡, statistical significantly different between chronic HCV versus DAA-cured liver slices. Levels of statistical significance *, P <0.05 **, P<0.01, ***, P<0.001. The mean and standard deviation within each group are plotted.

Figure S7

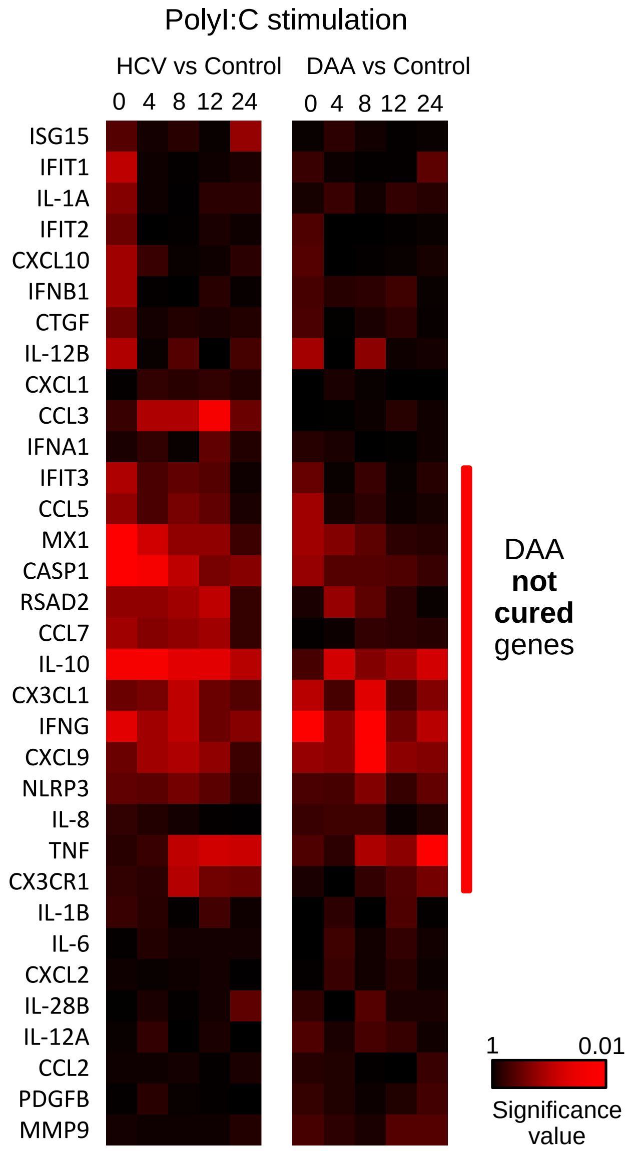


**Figure S7. Altered TLR3 response in chronic HCV-infected liver slices and DAA-cured liver slices.** (A) Hierarchical clustering of immune responsive genes with 2-fold or greater induction during poly-I:C stimulation. Genes and time points were colored based on the statistical significance P value of HCV-infected versus control, and DAA-cured versus control according to two-tailed Mann-Whitney test. The gene clusters that were altered by chronic HCV infection and not restored to normal by DAA therapy was highlighted.
