## Supplemental Table 1 for "Response of human liver tissue to innate immune stimuli"

Table S1  Patient information

**Chronic HCV-infected patients**

| Patient # | Patient ID | Diagnosis | HCV genotype | HCV RNA quantitation (IU/50 ng total RNA)[1] | | Prior HCV treatment |
|---|---|---|---|---|---|---|
| | | | | D0 | D7 | |
| 1 | Chronic HCV_1 | HCC | HCV G1a | 3371.3 | 2611.5 | Not treated |
| 2 | Chronic HCV_2 | ICC | HCV G1a | 8.2 | 1.8 | Not treated |
| 3 | Chronic HCV_3 | HCC | HCV G2 | 1.2 | 76.7 | Not treated |
| 4 | Chronic HCV_4 | HCC | HCV G1a | 3562.7 | 1762.0 | Not treated |
| 5 | Chronic HCV_5 | HCC | HCV G1b | 638.8 | 135.2 | Not treated |
| 6 | Chronic HCV_6 | HCC | HCV G1a | 3279.5 | 405.0 | Not treated |
| 7 | Chronic HCV_7 | HCC | HCV G1b | 9760.1 | 14365.8 | Not treated |
| 8 | Chronic HCV_8 | ICC | HCV G6 | 6769.8 | N/A | Not treated |

[1]The limit of detection in the assay was 1.2 IU/50 ng liver total RNA.

Table S1 (continued)

**HCV-DAA-cured patients**

| Patient # | Patient ID | Diagnosis | HCV genotype[1] | HCV RNA quantitation (IU/50ng total RNA) | DAA treatment duration | Time from completion of DAA treatment to HCC surgery with sample collection[3] |
|---|---|---|---|---|---|---|
| | | | | D0 | | |
| 1 | HCV-DAA_1 | HCC | HCV G1b | non-detect | 12 weeks | 9 months |
| 2 | HCV-DAA_2 | HCC | N/A | non-detect | N/A | N/A |
| 3 | HCV-DAA_3 | HCC | HCV G1a | non-detect | 12 weeks | 10 months |
| 4 | HCV-DAA_4 | HCC | HCV G1a | non-detect | 24 weeks | 12 months |
| 5 | HCV-DAA_5 | HCC | HCV G1b | non-detect | 12 weeks | 7 months |
| 6 | HCV-DAA_6 | HCC/ICC | N/A | non-detect | 8 weeks | less than 1 month |
| 7 | HCV-DAA_7 | ICC | HCV G1a or G1b | non-detect | 24 weeks | 9 months |
| 8 | HCV-DAA_8 | HCC | HCV G1a | non-detect | 12 weeks | less than 1 month |
| 9 | HCV-DAA_9 | HCC | HCV G1a | non-detect | 12 weeks | 20 months |
| 10 | HCV-DAA_10 | HCC | N/A | 0.3, Below LoD[2] | 12 weeks | 9 months |

[1]Genotype was obtained with clinical information prior to DAA treatment. N/A referred to HCV of unknown genotypes. Such patients were documented with HCV infection, however without genotype information.

[2]The 0.3 IU/50 ng liver total RNA was below the limit of detection (LoD), which was was 1.2 IU/50 ng liver total RNA. Thus, this patient was also HCV negative.

[3]This indicates the time from completion of DAA treatment to HCC surgery when the liver specimens were collected and analyzed.

Table S1 (continued)

**Non-infected patients**

| Patient # | Patient ID | Diagnosis | HCV RNA quantitation (IU/50ng total RNA) |
|---|---|---|---|
| | | | D0 |
| 1 | Non-infected_1 | Mullerian tumor | non-detect |
| 2 | Non-infected_2 | ICC | non-detect |
| 3 | Non-infected_3 | Cholangiocarcinoma | non-detect |
| 4 | Non-infected_4 | Colorectal cancer | non-detect |
| 5 | Non-infected_5 | Hepatocellular carcinoma | non-detect |
| 6 | Non-infected_6 | ICC | non-detect |
| 7 | Non-infected_7 | Neuroadenoma metastasis | non-detect |
| 8 | Non-infected_8 | CRC | non-detect |
| 9 | Non-infected_9 | Angiomyolipoma | non-detect |
